# Heritable confounding of exposure and outcome in Mendelian randomization studies

**DOI:** 10.1101/2024.09.05.24312293

**Authors:** Eleanor Sanderson, Dan Rosoff, Nicolai Vitt, Tom Palmer, Kate Tilling, George Davey Smith, Gibran Hemani

**Affiliations:** MRC Integrative Epidemiology Unit, University of Bristol, Bristol, UK; Population Health Sciences, University of Bristol, Bristol, UK; Section on Clinical Genomics and Experimental Therapeutics, National Institute on Alcohol Abuse and Alcoholism, National Institutes of Health, Bethesda, MD, USA; Oxford Centre for Diabetes, Endocrinology and Metabolism, University of Oxford, Oxford, UK

## Abstract

Mendelian randomization (MR) leverages genetic variants to infer causal effects of exposures on outcomes, assuming these variants influence outcomes solely through the exposure. However, genetic instruments associated with heritable confounders of the exposure and outcome can violate this assumption, undermining gene-environment equivalence and biasing MR effect estimates. With increasing sample sizes in genome-wide association studies genetic instruments with smaller effect sizes are being identified as associated with a trait. Here we use simulations and an applied example estimating the effect of C-reactive protein on type 2 diabetes to demonstrate that variants with smaller effect sizes are more prone to heritable confounding, leading to biased causal estimates across common MR methods. This bias acts in the same direction as the confounded associations between the exposure and outcome observed in linear regression, but often with greater magnitude and acts in the same direction across a number of commonly used MR estimation methods, potentially leading to misleading confidence in the results. We show that incorporating known or suspected heritable confounders via multivariable MR or applying Steiger filtering can mitigate this bias, which is sometimes not seen with methods aimed at dealing with correlated pleiotropy. These findings highlight the importance of assessing and adjusting for heritable confounding in MR analyses to improve causal inference reliability.

---

Epidemiological studies often aim to estimate causal effects from observational data. A common method is multivariable regression, which provides an unbiased estimate if all confounders of the exposure and outcome are known and measured without error, an assumption that is rarely plausible.[1] Mendelian Randomisation (MR) offers an alternative approach to causal effect estimation by using genetic variants as proxies for, or modifiers of, exposures to infer the causal effect of an exposure on an outcome, reducing bias from unmeasured confounding.[2–4] The fundamental principle of MR is of gene-environment equivalence[5], which requires biological reasoning that cannot be reduced to axioms. MR is currently most commonly implemented within an instrumental variable (IV) framework, using genetic variants associated with the exposure trait as instruments.[4]

IV analysis relies on three core assumptions (illustrated in Figure 1A to estimate the existence of a causal effect of the exposure on the outcome. An additional fourth assumption is required for interpretation of this estimate, (see [4] for an overview of the fourth IV assumption in the context of MR). In MR, single nucleotide polymorphisms (SNPs) identified as associated with the exposure in genome-wide association studies (GWAS) are generally employed as instruments. Typically, SNPs are selected if their association p-value with the exposure is <5×10⁻⁸, a Bonferroni-corrected threshold for genome-wide significance. They should therefore be considered to be putative instruments, although we use them as instrumental variables we are making the assumption that they satisfy the assumptions required to make them valid instruments. The reliability of the results obtained from a MR study depends on the putative genetic instruments satisfying the IV assumptions, however throughout the rest of the paper we will refer to genetic variants being used as instrumental variables as instruments even when those assumptions don’t hold as that is how they are used in a naïve MR study.[6]

**Figure 1:**
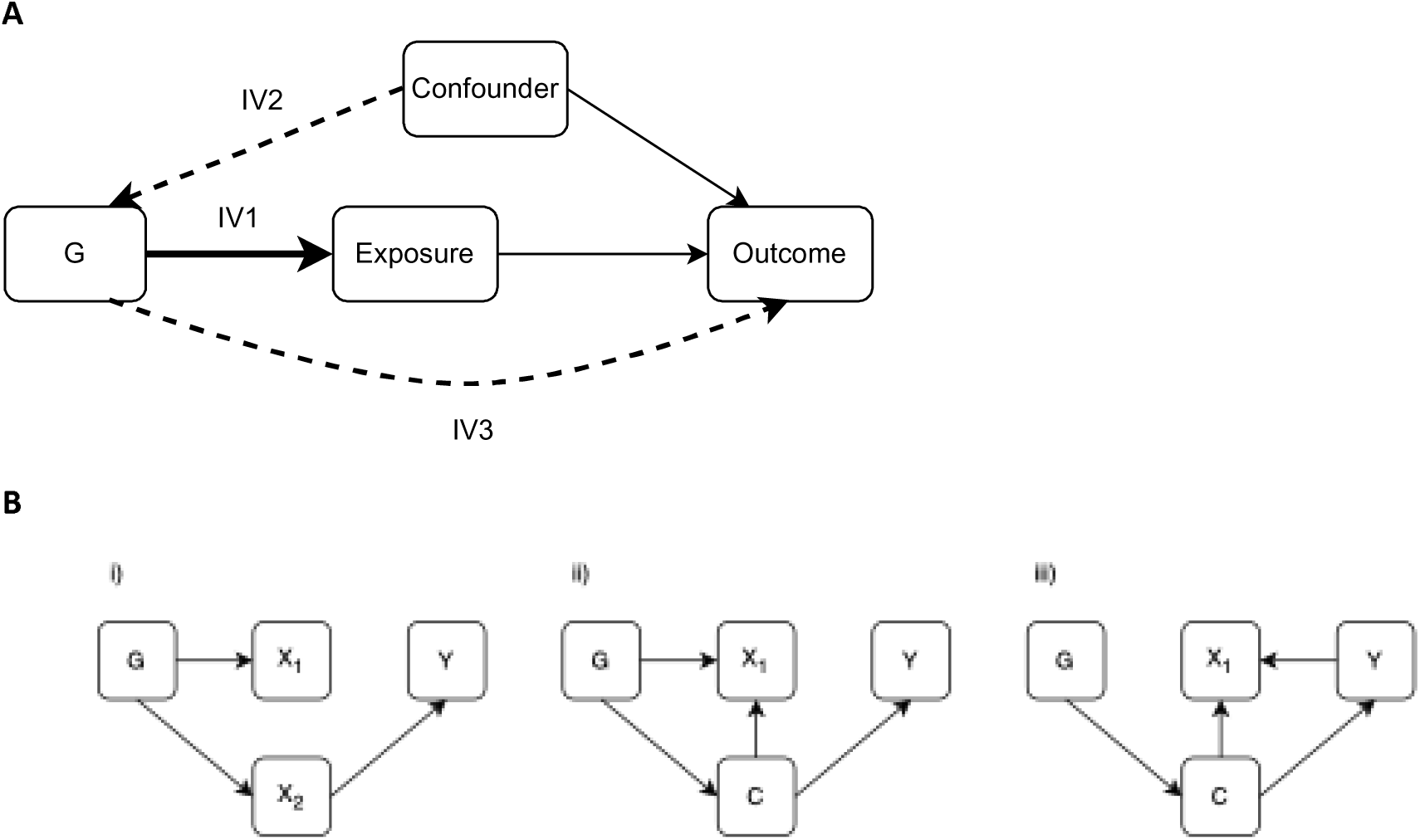
Instrumental Variable assumptions and potential mechanisms that can generate correlated pleiotropy and bias MR. Figure illustrating the IV assumptions and the different mechanisms that can cause correlated pleiotropy in MR studies in the absence of a causal effect of the exposure on the outcome. X is the exposure of interest, Y is the outcome, C is a confounder of the exposure and outcome. G is a set of SNPs associated with the exposure, some of which may be associated through another phenotype, X_2_ is a second exposure that is not the exposure of interest. A. Illustration of the main IV assumptions; (1) the IV should be associated with the exposure, (2) there should be no confounding of the IV and the outcome and (3) there should be no pathway from the IV to the outcome that doesn’t act through the exposure. The dashed lines represent associations that should not exist for the IV assumptions to be satisfied. B. i) Uncorrelated horizontal pleiotropy, some of the SNPs in G are associated with Y through X_2_ ii) heritable confounding, some of the SNPs in G are associated with Y through a confounder of X_1_ and Y iii) reverse causation, the SNPs in G are associated with Y through a confounder of X_1_ and Y and the direction of effect has been mis-specified.

The polygenic nature of complex traits, for which there is substantial empirical support [7], posits that for most traits the majority of heritable variation is driven by a very large number of genetic factors, each with very small effect sizes. To discover such variants GWAS have been growing in sample size rapidly in recent years, especially through the emergence of large scale biobanks. For example, a recent GWAS of height meta-analysed data from 5.4 million individuals.[8] Correspondingly, increasing numbers of variants are being identified as associated with the traits studied in these GWAS.

## The structure of “heritable confounding”

For MR analyses involving exposures that are complex traits, the effects of genetic variants associated with the exposure on that exposure are mediated through several phenotypic layers.[9] Germline genetic variation influences the traits that are generally instrumented in MR through a wide variety of phenotypic factors: gene expression levels, mRNA splicing, DNA methylation, protein abundance, post-translational protein modifications, enzyme activity, etc.[10] Such mediation opens the door for genetic variants to act on multiple different pathways if these mediating factors have effects on multiple downstream traits. This can bias MR estimates by introducing horizontal pleiotropy, violating assumption IV3, if there is a path from the genetic variant to the outcome that does not act via the exposure (see **Figure 1A**).

Correlated pleiotropy is used to describe a scenario where the horizontal pleiotropic effect is correlated with the magnitude of association between the genetic variants and the exposure. A heritable confounder can generate correlated pleiotropy, as illustrated in **Figure 1B**. Heritable confounding occurs when the genetic variants used as instruments are associated with a phenotypic confounder of the exposure and outcome, leading to a violation of the exclusion restriction (IV3).

This process can lead to an investigator using a genetic variant that primarily influences a heritable confounder to be an instrument for a phenotype downstream of that heritable confounder.[11] This problem has been referred to as misspecification of the primary phenotype in the MR literature.[3, 4] but here we will use the term heritable confounding to refer to any situations where some of the genetic instruments used are associated with a confounder of the exposure and outcome.

Confounding of the instrument and outcome is an alternative source of bias in MR studies that can occur due to population stratification, dynastic effects and assortative mating. This will lead to a violation in the second IV assumption (independence). There is a separate literature on this bias and how to correct for it.[12, 13] Confounding of the instrument and the outcome is not the focus of this paper, we therefore use the term heritable confounder to refer to a phenotypic confounder of the exposure and outcome that is influenced by genetic variation throughout.

Reverse causation can occur in MR when variants associated with either the outcome itself (e.g. myocardial infarction) or the disease process leading up to the outcome (e.g. atherosclerosis), are included as instruments for the exposure. This can also lead to heritable confounding when the disease process that leads to the reverse causation is a confounder of the exposure and the outcome, illustrated in **Figure 1B**.

While heritable confounding is a potential source of bias in all MR analyses, many MR studies currently do not explore the potential for this type of bias. In this paper we consider the consequences of heritable confounding for MR studies and propose two alternative applications of currently used approaches to identify and correct for bias from heritable confounding.

Alternative MR methods have been developed to help identify and/or adjust for correlated pleiotropy under specific assumptions about its structure.[14–24] These approaches are often computationally intensive, using Bayesian estimation approaches applied to a large number of genetic variants. They also impose an assumption of a structure of the relationship between the pleiotropic variants and other variants, often assuming that the plurality of the instruments are valid. The approaches we propose here are already widely used in other settings and are simple to apply, leveraging empirical evidence on pleiotropic relationships that are openly accessible in large scale databases. We therefore propose that they should be systematically used within MR studies to identify how sensitive the results obtained are to the most likely confounders.[5]

The rest of the paper is structured as follows. Firstly, we compare the bias in MR studies to the bias from residual confounding in linear regression, showing that bias in MR estimation from a single heritable confounder can exceed the bias from the same confounder in multivariable or even univariable regression. We then investigate whether increasing GWAS sample sizes could increase the rate of correlated pleiotropy through a simulation of a network of traits explained by a set of genetic variants. We go on to evaluate how this increase in correlated pleiotropy impacts standard MR estimators in a series of simulations. In settings where potential confounders are hypothesised and measurable, we propose two approaches to mitigate bias: (1) multivariable MR (MVMR)[25], including the confounder as an additional exposure, and (2) Steiger filtering, removing SNPs that explain more variance in the confounder than the exposure.[26] To illustrate the potential use of these approaches we apply both methods, along with existing approaches, to estimate the effect of C-reactive protein (CRP) on type 2 diabetes, using phenome-wide association studies (PheWAS) to identify potential heritable confounders.

## Bias in IVW MR estimates compared to linear regression

In this section we explore the bias of MR compared to the equivalent linear regression in the presence of a single unmeasured heritable confounder. We consider a simple model of a relationship between an exposure (*X*) and an outcome (*Y*) which is confounded by a single unobserved heritable confounder (*U*). The exposure is associated with a single instrument (*Z*) that also has an effect on the outcome via the unobserved confounder. We assume that all relationships are linear and there are no interactions. We focus on the setting with a single heritable confounder for simplicity and clarity; the results given here therefore only compare the results for bias from a single heritable confounder and not the overall bias that would be expected. The model we consider can be written as;

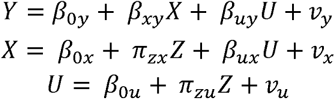

Where *v_y_, v_x_* and *v_u_* are all uncorrelated normally distributed random error terms. This model is illustrated in **Figure 2A**.

**Figure 2.**
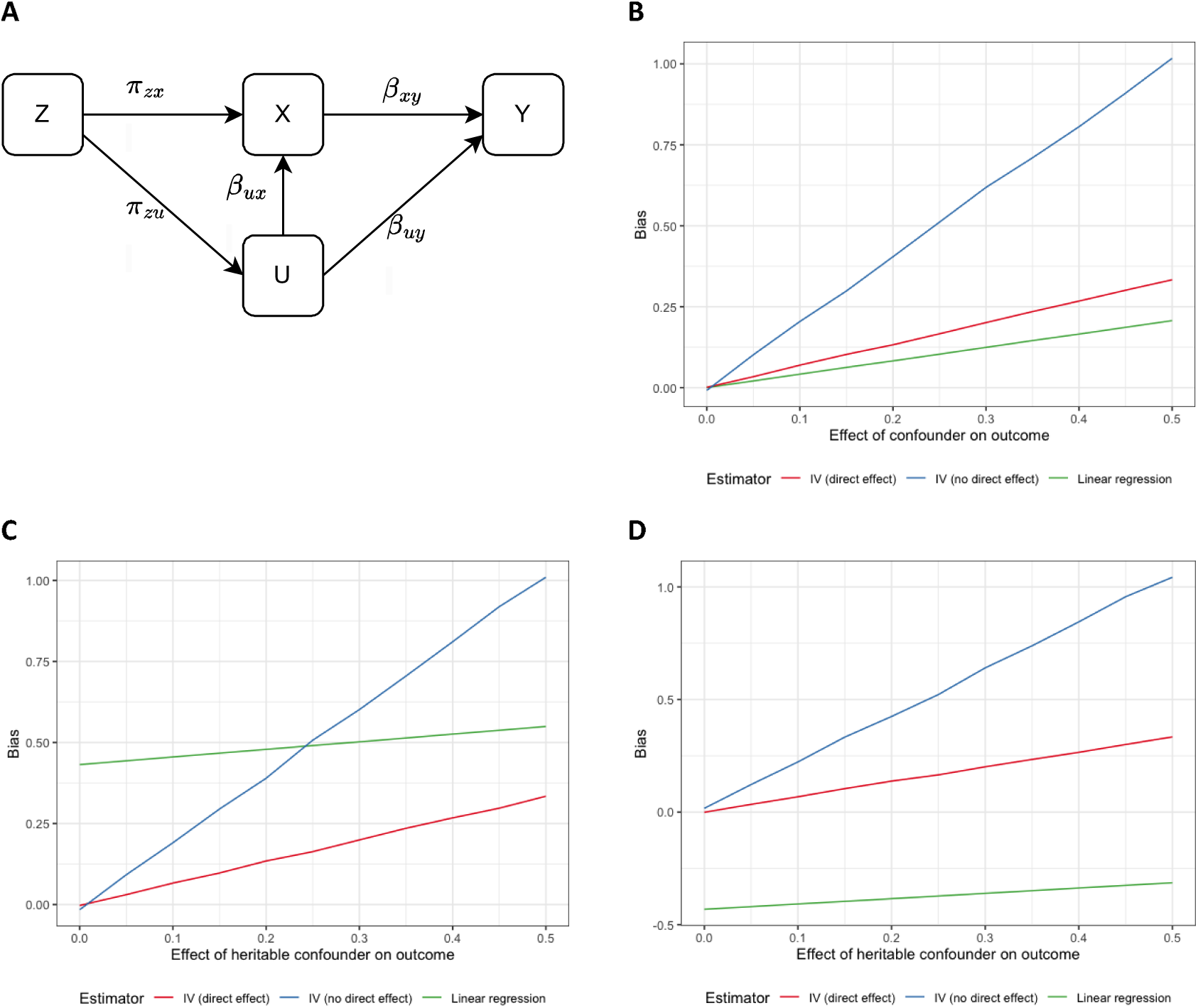
The Bias of linear regression and IV estimators when instruments have a pleiotropic effect through a single heritable confounder. A. A simple model with a single IV associated with an exposure and a confounder. This is used in obtaining the expressions for bias given above and for generating the simulation results in (B). B. Bias from estimation by linear regression and two-stage least squares for the model in (A) estimated assuming U is unobserved and there is no other confounding. Linear Regression gives the bias of the observational association, IV (direct effect) sets and gives the bias of the IV estimator, IV (no direct effect) sets and gives the bias of the IV estimator. The relative positions of these plots depends on the other parameters in the model and the assumption of no other (non-heritable) confounding. C. Bias plots given in B with the addition of a positive non-heritable confounder of the exposure and outcome. D. Bias plots given in B with the addition of a negative non-heritable confounder of the exposure and outcome.

We show in **Supplementary material section 1.1** that the bias in the MR estimate from a single heritable confounder will be less than the bias of the linear regression estimate from the same confounder when;

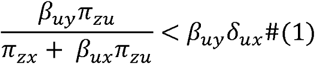

Where *δ_ux_* is the association between *U* and *X*, estimated from an unadjusted regression of *X* on U. The implication of equation (1) is that the bias depends on the ratio of the pleiotropic effect of the instrument on the outcome (*β_uy_π_zu_*) to the effect of the instrument on the exposure (*π_zx_ + β_ux_ π_zu_*). The larger the pleiotropic effect relative to the effect of the instrument on the exposure the larger the bias the MR estimator will be. As the association between the variant and the exposure *π_zx_* increases the overall bias of the MR estimation will decrease.

To illustrate the relationship between the bias of linear regression and IV estimation across different levels of pleiotropy we simulated the model given in **Figure 2A** varying the effect of the instrument on the confounder but holding all other parameters constant. The mean bias of each estimator across 1000 repetitions for varying levels of confounding is given in **Figure 2B**. These results illustrate how the bias of the IV estimator increases as the pleiotropic effect via the confounder increases and can be substantially more than the bias in the purely observational linear regression. The exact position and slope of these lines will depend on the other parameters in the model.

**Figures 2C** and **2D** show the same bias plots as **Figure 2B** with an additional non-heritable confounder added to the model. The direction of the confounding from this additional confounder is varied from positive (**Figure 2C**) to negative (**Figure 2D**). These plots highlight how the overall bias of each estimator will depend on other factors as well as the level of heritable confounding and the bias of the estimate from MR may not always act in the same direction as the bias of the estimate from linear regression.

## Network simulations

Having described the impact of correlated pleiotropy on MR performance, we now illustrate that under a basic model of a random genotype-phenotype map, we would expect a relationship between smaller instrument-exposure effect size and higher probability of acting via a heritable confounder. Hence we hypothesise that as GWAS sample sizes increase, MR studies will become increasingly vulnerable to horizontal pleiotropy because variants with smaller effects on the exposure will increasingly be included as instruments. One reason that a variant may have a small marginal effect on the exposure is that its influence is transmitted through a relatively distal biological pathway involving multiple mediating traits. In such cases the overall effect is attenuated by the product of the path coefficients along the causal chain. In a random genotype–phenotype map, variants that reach the exposure through more distal pathways are also expected to be more phenotypically connected overall. Consequently, if any upstream mediating trait also influences the outcome through a pathway that does not pass through the exposure, the variant will induce heritable confounding in the MR estimate.[27] This model is illustrated in **Figure 3a**, where the variants with the shortest path to the exposure do not lead to correlated pleiotropy but the variant with the longest path to the exposure does.

**Figure 3.**
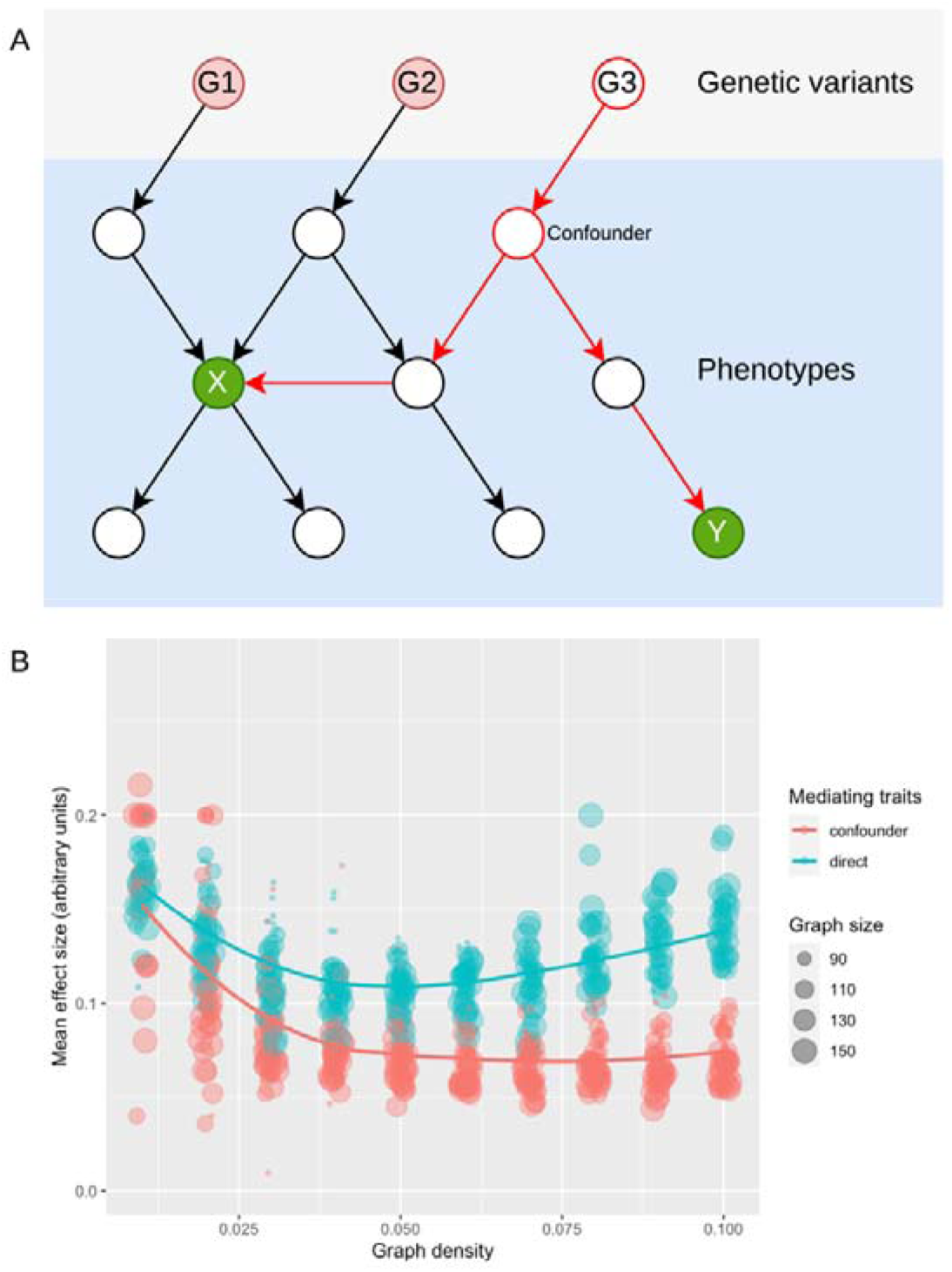
Network-based simulations of the origin of instruments. A. Illustration of a random network. For each possible trait pair we assign an exposure (X) and an outcome (Y). In this simplified example instrument G1 is considered to act ‘directly’ because there is no confounding path between X and Y, G2 may exhibit greater pleiotropy but without inducing bias as it has no path to Y that is not through X, whereas G3 is considered to act via a confounding path, and by being more distal from X it has a greater chance of finding a path to Y. The effect size of each instrument is determined based on all edges having a uniform effect of 0.2. B. The mean effect of the instrument on the exposure (y-axis) across simulations (x-axis and point size), stratified by whether they act ‘directly’ or via confounding paths. We observe that in general direct instruments have larger effects than those acting via confounders.

We examined this relationship through simulations. We use the randomDAG function in the R/dagitty package to simulate a DAG with an arbitrary number of nodes, and a fixed probability that any set of nodes is connected. Nodes with no parents in the simulated DAG are identified as genetic variants, and 500 trait pairs are selected at random, in each pair assigning one to be the hypothesised exposure and one to be the hypothesised outcome. We then identify the path length for every genetic variant for the exposure, and also determine whether they act via a confounder, or otherwise (i.e. directly or through other traits that mediate the association between the genetic variant and the exposure and have no other paths to the outcome). The effect size of the genetic variant on the exposure is *β^L^* where *L* is the path length from genotype to exposure and, for simplicity, every edge in the DAG has the same effect *β* = 0.2. These simulations were conducted for a set of parameters of graphs with number of traits N=75, 100, 125, 150 and graph density based on probability of an edge occurring ranging from p = 0.01, 0.02,…, 0.1. Each parameter combination was repeated 10 times.

The results from this simulation are given in **Figure 3b, illustrating** that instruments that act via confounders tend to have smaller effects due to more traits lying on the path from the genetic variant to the exposure. While this simulation is designed around a baseline model to motivate the potential importance of heritable confounding, the architecture of a genotype-phenotype map may give rise to other relationships between instrument effect size and correlated pleiotropy. Particularly we have assumed that all edges have the same magnitude of effect and that the distribution of the connections is random. In any case it is important to consider empirically potential signatures of heritable confounding in MR analyses.

## Simulations evaluating standard MR methods under heritable confounding

We next simulated a model where the exposure and outcome are confounded by a third, potentially unobserved, heritable trait. In this model, the exposure has no causal effect on the outcome. The exposure is influenced by two upstream traits: one is a mediator of the effect of the genetic variant on the outcome, which simply transmits genetic effects to the exposure, while the other is a heritable confounder affecting both exposure and outcome. Note that if a different outcome were being studied it is possible that the first upstream trait mentioned above is the one that generates bias due to horizontal pleiotropy, and the second upstream trait that does not do so.

Linear regression of the outcome on the exposure gives a positive estimate of the association between the exposure and outcome due to the existence of a confounder that affects both in the same direction. The mediator and confounder are both influenced by a separate set of genetic variants. We assume no non-heritable confounding and that all instrumental variable (IV) assumptions otherwise hold. The model corresponds to **Figure 1B(ii)** and is illustrated in **Figure 4**.

**Figure 4.**
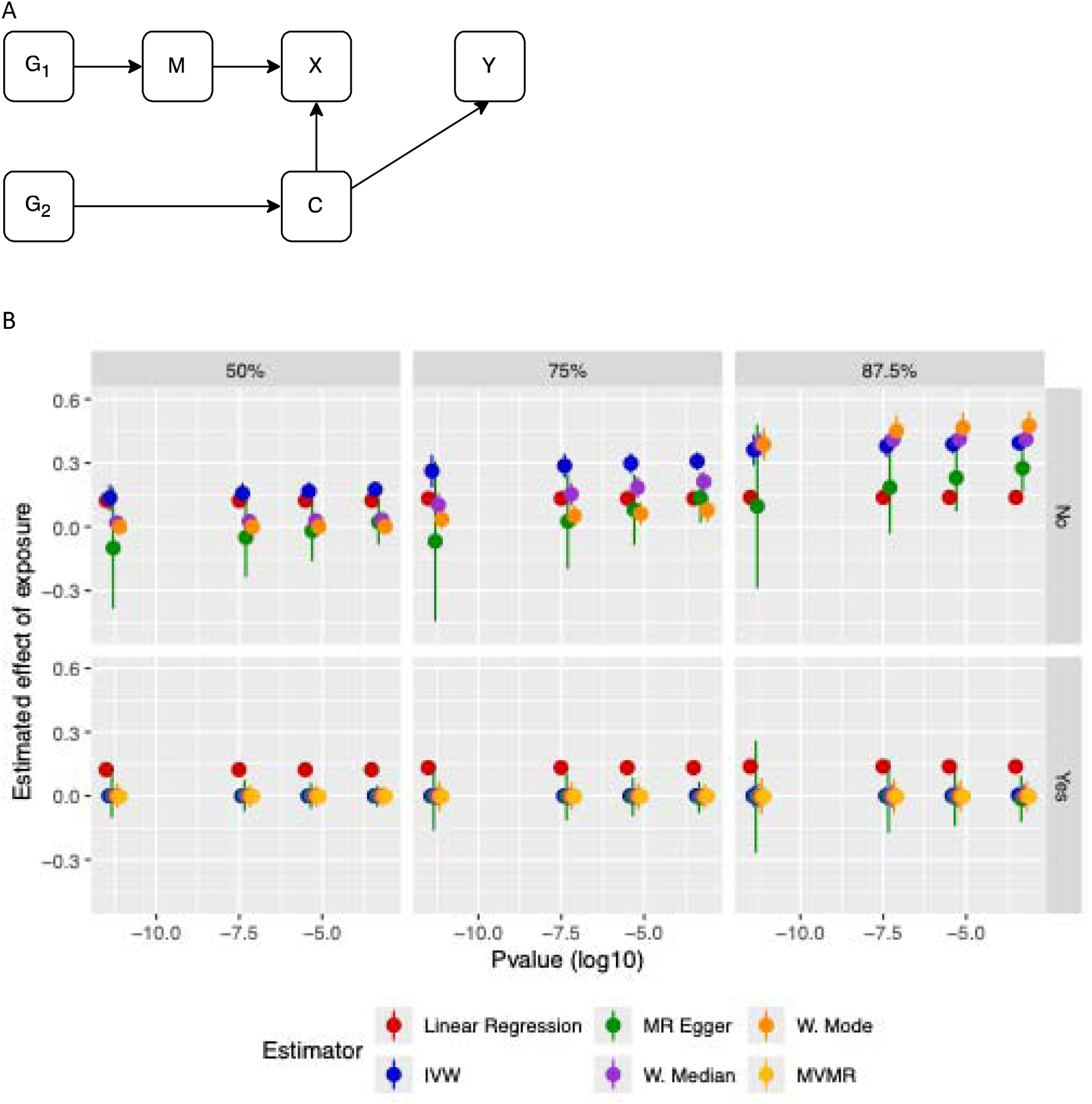
Simulation of bias from a single heritable confounder in linear regression and MR. A. The model used to generate the data in the simulation. X is the exposure of interest, Y is the outcome, C is a (potentially observable) confounder of the exposure and outcome, M is a mediator upstream of X. G1 is a set of SNPs associated with the mediator, G2 is a set of SNPs associated with the confounder. SNPs in G1 may be associated with the exposure through the mediator and SNPs in G2 may be associated with the exposure through the confounder. There is no causal effect of the exposure on the outcome. B. Results for the simulations varying; the proportion of the SNPs in G2 (50%, 75% or 87.5%), whether confounder adjustment (Steiger filtering/MVMR) is applied (No/Yes) and the p-value threshold applied (given on X-axis) to illustrate different GWAS sample sizes. Total number of SNPs – 400, % given is the % in G2, the rest are valid SNPs for X (i.e. G1). All SNPs that are significantly associated with X in the simulated data are included in the univariable MR estimation and all SNPs significantly associated with X or C are included in the MVMR estimation. There is no causal effect of X on Y, the effect of M on X is 1, of C on Y is 0.4 and of C on X is 0.75. n=10000, reps=1000. The effect of X on Y is estimated by; Linear regression, IVW, MR Egger, weighted median, weighted mode and MVMR including C as an additional exposure. Steiger filtered results includes all univariable estimators with Steiger filtering applied to remove SNPs that explain more variation in C than X.

We simulated two independent samples of 100,000 individuals each. In each sample, 400 SNPs were generated from a binomial distribution with the proportion of SNPs associated with the mediator and confounder varying across the simulations—ranging from a 50% associated with each to 12.5% associated with the mediator and 87.5% associated with the confounder.

SNP effects on the mediator or confounder were drawn from a normal distribution (mean = 0, SD = 0.04), orientated so all SNPs had a positive association with the relevant trait. The confounder and mediator were generated as the sum of their respective SNP effects plus a normally distributed error term. The exposure was generated as a linear combination of the mediator, confounder, and an error term. As shown earlier, SNPs associated with a mediator may be more strongly associated with the exposure than SNPs associated with a confounder. Therefore, we down-weighted the confounder – exposure association by 25% relative to the mediator – exposure association to mimic this more distant association. Results with this down weighting varied to 0% and 50% are given in the **Supplementary Figure 1** and **Supplementary Figure 2** respectively. Finally, the outcome was generated as the effect of the confounder and a random error term. The expressions for the data generation process are given in **Supplementary material section 1.2**.

GWAS summary statistics were generated by regressing each of the exposure and confounder on each SNP separately in Sample 1, and the outcome on each SNP in Sample 2. SNPs (from either set) were selected to be used as instruments in our analysis based on their association with the exposure in Sample 1. Increasing the sample size of a GWAS will decrease the standard error in the SNP-trait association proportionally across all SNPs leading to more SNPs being selected based on a fixed p-value threshold cut-off. Therefore, to model varying the sample size in the GWAS while keeping all other aspects of the data constant we varied the p-value threshold used to select SNPs for use as instruments in the MR across our simulations with larger p-values being equivalent to a larger sample size. The same sample was used for SNP selection and exposure effect estimation, reflecting standard MR practice.

Using the summary statistics generated we applied four common univariable MR methods; Inverse Variance Weighted (IVW)[28], MR Egger[29], Weighted Median[30], and Weighted Mode[31]. We also applied Multivariable MR (MVMR)[25, 32], including the confounder as an additional exposure and SNPs associated with the confounder as additional instruments. In addition, we repeated all univariable MR methods with Steiger filtering[26] applied to exclude SNPs that explained more variation in the confounder than in the exposure.

MVMR is a method that allows for multiple exposures to be included in a single MR estimation, with all genetic variants associated with any of the exposures included as instruments.[25] Effect estimates obtained from MVMR should be interpreted as the direct effect of each exposure on the outcome conditional on the other exposures included in the analysis. If the additional exposure is a confounder of the exposure and outcome this will not change the interpretation from a univariable MR estimation, however caution needs to be applied with the inclusion of potential mediators of the exposure and outcome as the estimand targeted will differ between a univariable MR and MVMR estimation.[25]

Steiger filtering is usually used as an approach to identify individual genetic variants that explain more variation in the outcome than the exposure. It works by comparing the proportion of the variation explained (accounting for sample size) in each of the exposure and outcome by an individual variant. It can then be used to remove variants that explain more variation in the outcome than the exposure (at p<0.05).[26] Here we adapt the method to identify genetic variants that likely associate with the exposure acting primarily through a potential confounder. This is achieved by comparing the proportion of the variation explained by each a variant in the exposure and the identified potential heritable confounders, removing those as instruments that explain more variance in a confounder. There are limitations in the application of the method with binary traits where R^2^ is estimated on the liability scale. Additionally, the requirement for the variant to explain statistically significantly more variation in the confounder/outcome than the exposure means that variants may be erroneously retained in the analysis due to uncertainty in the variation explained or the proportion of variation explained is very similar between each trait.

Results from this simulation are given in **Figure 4** and **Supplementary Tables 1 and 2**. When the total proportion of the variation in the exposure explained by genetic variants that act on that exposure through the confounder is moderate IVW and MR Egger are biased across all p-value selection thresholds. However both weighted median and weighted mode correctly estimate the causal effect of the exposure on the outcome as their assumption about the nature of any pleiotropy is still satisfied (**Figure 4, Supplementary Table 1**). When almost all of the SNPs are associated with the confounder all univariable methods of estimation are biased and give results that are consistent with each other (**Figure 4, Supplementary Table 1**). With a stricter p-value cut-off, equivalent to a smaller sample size, MR Egger is very imprecise. This is expected as a stricter p-value cut off will mean fewer SNPs are included in the estimation and MR Egger has low power when the number of SNPs included in the estimation is small. In all cases both MVMR and the univariable MR approaches with Steiger filtering between the exposure and confounder applied gave unbiased estimates of the causal effect of the exposure on the outcome. However, each of these methods requires the confounder to be known and have available GWAS summary statistics.

**Table 1** shows the mean number of genetic variants in each simulation that were selected for the univariable MR estimation methods (IVW, MR Egger, weighted median and weighted mode). When the mediator is associated with as many SNPs as the confounder in the data generating process the final proportion of SNPs that are associated with the confounder in the MR estimation is small due to the effects of those SNPs on the exposure being smaller.

**Table 1.** The number of SNPs selected in each simulation.

| *P-value cut off* | *No. SNPs* | *Exposure* | *Confounder* | *% for confounder* |
| --- | --- | --- | --- | --- |
| **50% of simulated SNPs associated with confounder** |  |  |  |  |
| $5 \times 10^{-12}$ | 57 | 39 | 18 | 31% |
| $5 \times 10^{-8}$ | 97 | 61 | 36 | 37% |
| $5 \times 10^{-6}$ | 131 | 79 | 52 | 40% |
| $5 \times 10^{-4}$ | 182 | 104 | 78 | 43% |
| **87.5% of simulated SNPs associated with confounder** |  |  |  |  |
| $5 \times 10^{-12}$ | 42 | 10 | 32 | 76% |
| $5 \times 10^{-8}$ | 79 | 15 | 64 | 80% |
| $5 \times 10^{-6}$ | 112 | 20 | 92 | 82% |
| $5 \times 10^{-4}$ | 164 | 27 | 137 | 84% |

The number of SNPs selected as associated with the exposure (out of the 400 generated) based on the p-value cutoff used for selection. Exposure gives the number of those that were generated as associated with the exposure and Confounder the number of those that were generated as associated with the confounder but selected as associated with the exposure. Numbers are means across all iterations, rounded to the nearest whole number.

In these simulations there is a single confounder of the exposure and outcome of interest biasing the estimates obtained. These results therefore show how the MR effect estimates are biased by that confounder in the same direction as the confounded observational association and may be substantially more biased than the observational association. In practice it is likely that there may be many potential confounders, some of which will be heritable and associated with a subset of the genetic variants selected as instruments. The overall bias in each method will depend on a number of other factors (Linear regression; further confounding, measurement error, selection bias etc. MR; weak instruments, population structure, further heritable confounders, selection bias etc.). These results therefore do not show the overall bias in either type of estimation. With multiple potential heritable confounders both Steiger filtering and MVMR could be applied to each of them in turn to identify any bias. We illustrate this approach in the application below.

## Illustrative application: Estimation of the effect of C-Reactive protein on Type 2 Diabetes

In our illustrative application we considered a potential exposure that has been widely used in MR studies; C-Reactive Protein (CRP), a marker of inflammation and estimate its effect on Type 2 Diabetes (T2D), exploring the possibility of heritable confounding biasing the observed effects.[33–36] A recent GWAS comprising >500,000 individuals with European genetic ancestry[37] identified 728 lead variants (P < 5×10^-8^; LD r^2^ < 0.1) across 266 loci associated with CRP, many of which may be associated due to their influence on upstream traits which influence outcomes independent of CRP, and thus be potential heritable confounders. We use MR to estimate the effect of CRP on T2D and systematically identify potential confounders and remove SNPs more strongly associated with those confounders than CRP. We compare the results obtained to a set of alternative MR methods that may identify confounding.

To identify traits associated with the putative CRP instruments, we used the FUMA platform[38] and performed a trait look-up in the GWAS Catalog (accessed 10^th^ April 2024). This yielded 33,573 SNP–trait associations (P < 5×10^-8^) among the lead SNPs or variants in strong LD (r^2^ > 0.6). From this, we curated a set of 5,198 traits for further analysis, prioritizing biomarkers and risk factors over disease outcomes.

To identify which of these traits are potential confounders we applied summary data MR to estimate the effect of each trait identified on firstly CRP and then T2D. Traits showing nominal effects on both CRP and T2D were retained as potential heritable confounders.

To estimate the effect of each trait on CRP we selected genome wide significant variants for each trait (P < 5×10⁻⁸; r^2^ < 0.001) as instruments using data from the MRC IEU OpenGWAS platform.[39] When only one variant was available for a trait, we used the Wald ratio; for traits with two or more variants, we applied inverse-variance weighted (IVW) MR; and for traits with three or more variants, we also included MR Egger[29], Weighted Median[30], and Weighted Mode[31] estimates. **Table 2** summarises the proportion of the analyses conducted that indicated evidence of a causal effect using a heuristic threshold (p < 0.05) to identify associations that indicate a potential causal effect. Depending on the MR method applied, 20–30% of traits with available instruments showed nominal evidence of an effect on CRP.

**Table 2.** Proportion of traits that show evidence of a causal effect on C-Reactive Protein in MR.

|  | **No of tests** | **% P < 0.05** | **% P < 0.01** |
| --- | --- | --- | --- |
| *Wald ratio/IVW* | 3770 | 21.7% | 13.9% |
| *MR Egger* | 1480 | 27.5% | 19.2% |
| *Weighted median* | 1478 | 29.0% | 19.6% |
| *Weighted mode* | 1478 | 22.7% | 13.7% |

Results from MR estimation of the effect of up to 3770 traits on CRP. Traits were selected from those strongly associated with at least one lead SNP in a recent CRP GWAS. Wald ratio was used for traits with 1 SNP available as an instrument and IVW was used for traits with 2 or more SNPs available as instruments. MR Egger, Weighted median and Weighted mode were additionally applied for all traits with 3 or more SNPs available as instruments.

865 traits showing putative evidence of an effect on CRP (p < 0.01 using Wald ratio or IVW) were subsequently tested for their effect on T2D using the same process as for CRP. Traits which showed evidence of an effect on T2D (p < 0.01 using Wald ratio or IVW) were retained as potential heritable confounders. This resulted in 235 potential heritable confounders. We have not corrected for multiple testing or the potential bias in this stage of the analysis to maximise the number of potentially heritable confounding traits taken forward. However bias in the MR estimates will have led to a number of false positives and similarly low power will have led to some false negatives, not identifying true heritable confounders.

We estimated the causal effect of CRP on T2D using summary data MR with all genome-wide significant CRP variants (P < 5×10⁻⁸; r^2^ < 0.001). If the SNPs used as instruments for CRP are also associated with T2D through a heritable confounder this MR estimate will be biased. Therefore to identify any bias from heritable confounding, we applied Steiger filtering, removing variants that explained more variance in each potential heritable confounder identified above than in CRP (p < 0.05) in turn. Where these potential heritable confounders are not continuous this variance is measured on the liability scale of the exposures. Variants that explain more variation in the potential heritable confounder than in CRP are likely to be associated with CRP through that potential heritable confounder and so have a horizontally pleiotropic effect on T2D through the confounder, biasing the MR effect estimate of CRP on T2D.

**Table 3** gives results for the 10 confounders that had the largest effect on the MR estimate of CRP on T2D obtained (measured by IVW p-value). For these 10 confounders we also applied MVMR to estimate the effect of CRP on T2D controlling for each confounder in turn. In each model we estimated the conditional F-statistics to test instrument strength.[40] **Table 3** gives the estimated effect for CRP adjusted for each confounder where conditional F-statistics for both exposures were greater than 10, suggesting sufficient instrument strength for estimation. Full results are provided in **Supplementary Section 2**.

**Table 3.**
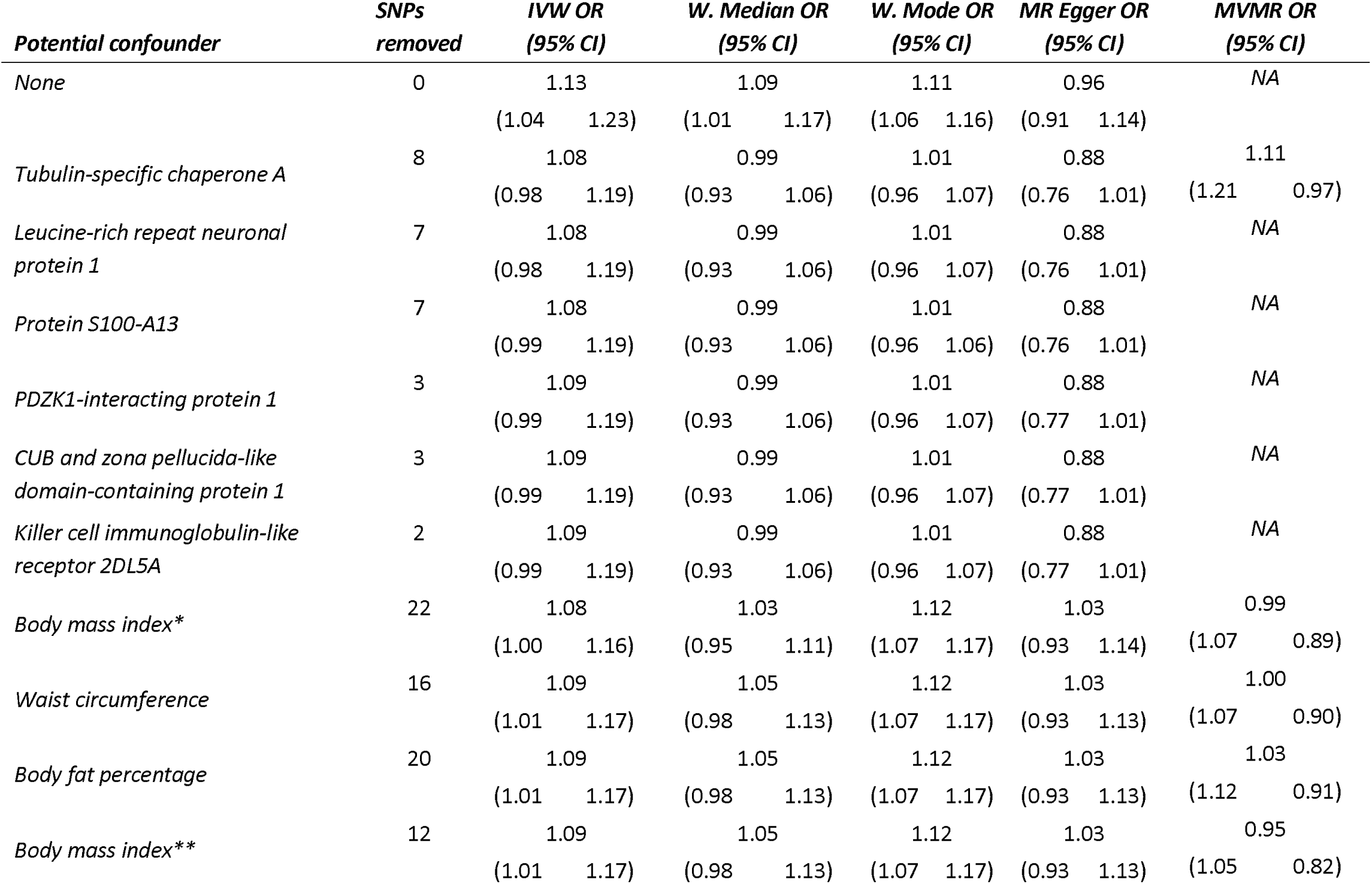
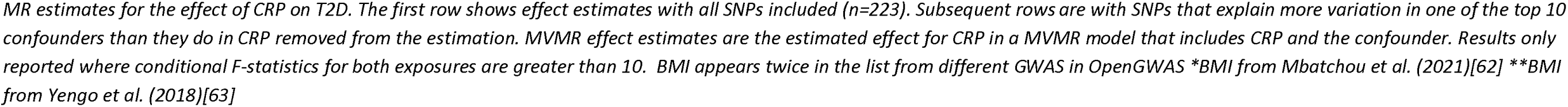
MR effect estimates for CRP on T2D, with all SNPs included and with SNPs more strongly associated with each of the top 10 confounders removed.

| *Potential confounder* | *SNPs removed* | *IVW OR (95% CI)* | *W. Median OR (95% CI)* | *W. Mode OR (95% CI)* | *MR Egger OR (95% CI)* | *MVMR OR (95% CI)* |
| --- | --- | --- | --- | --- | --- | --- |
| *None* | 0 | 1.13<br>(1.04 1.23) | 1.09<br>(1.01 1.17) | 1.11<br>(1.06 1.16) | 0.96<br>(0.91 1.14) | NA |
| *Tubulin-specific chaperone A* | 8 | 1.08<br>(0.98 1.19) | 0.99<br>(0.93 1.06) | 1.01<br>(0.96 1.07) | 0.88<br>(0.76 1.01) | 1.11<br>(1.21 0.97) |
| *Leucine-rich repeat neuronal protein 1* | 7 | 1.08<br>(0.98 1.19) | 0.99<br>(0.93 1.06) | 1.01<br>(0.96 1.07) | 0.88<br>(0.76 1.01) | NA |
| *Protein S100-A13* | 7 | 1.08<br>(0.99 1.19) | 0.99<br>(0.93 1.06) | 1.01<br>(0.96 1.06) | 0.88<br>(0.76 1.01) | NA |
| *PDZK1-interacting protein 1* | 3 | 1.09<br>(0.99 1.19) | 0.99<br>(0.93 1.06) | 1.01<br>(0.96 1.07) | 0.88<br>(0.77 1.01) | NA |
| *CUB and zona pellucida-like domain-containing protein 1* | 3 | 1.09<br>(0.99 1.19) | 0.99<br>(0.93 1.06) | 1.01<br>(0.96 1.07) | 0.88<br>(0.77 1.01) | NA |
| *Killer cell immunoglobulin-like receptor 2DL5A* | 2 | 1.09<br>(0.99 1.19) | 0.99<br>(0.93 1.06) | 1.01<br>(0.96 1.07) | 0.88<br>(0.77 1.01) | NA |
| *Body mass index\** | 22 | 1.08<br>(1.00 1.16) | 1.03<br>(0.95 1.11) | 1.12<br>(1.07 1.17) | 1.03<br>(0.93 1.14) | 0.99<br>(1.07 0.89) |
| *Waist circumference* | 16 | 1.09<br>(1.01 1.17) | 1.05<br>(0.98 1.13) | 1.12<br>(1.07 1.17) | 1.03<br>(0.93 1.13) | 1.00<br>(1.07 0.90) |
| *Body fat percentage* | 20 | 1.09<br>(1.01 1.17) | 1.05<br>(0.98 1.13) | 1.12<br>(1.07 1.17) | 1.03<br>(0.93 1.13) | 1.03<br>(1.12 0.91) |
| *Body mass index\*\** | 12 | 1.09<br>(1.01 1.17) | 1.05<br>(0.98 1.13) | 1.12<br>(1.07 1.17) | 1.03<br>(0.93 1.13) | 0.95<br>(1.05 0.82) |

*MR estimates for the effect of CRP on T2D. The first row shows effect estimates with all SNPs included (n=223). Subsequent rows are with SNPs that explain more variation in one of the top 10 confounders than they do in CRP removed from the estimation. MVMR effect estimates are the estimated effect for CRP in a MVMR model that includes CRP and the confounder. Results only reported where conditional F-statistics for both exposures are greater than 10. BMI appears twice in the list from different GWAS in OpenGWAS \*BMI from Mbatchou et al. (2021)[62] \*\*BMI from Yengo et al. (2018)[63]*

**Figure 5a** shows a scatterplot of the variants used in the CRP–T2D MR analysis, with those associated with the top 10 confounders highlighted. These confounders clustered into two major groups: adiposity-related traits and protein biomarkers. The consistent appearance of adiposity-related traits suggests adiposity is a key confounder in the CRP–T2D relationship. Excluding variants associated with these traits led to substantial attenuation of the MR effect estimate for CRP on T2D, reducing evidence for a causal relationship. MVMR analysis adjusting for adiposity traits also led to a substantial attenuation of the effect estimate with all analyses compatible with a null causal effect of CRP on T2D.

**Figure 5.**
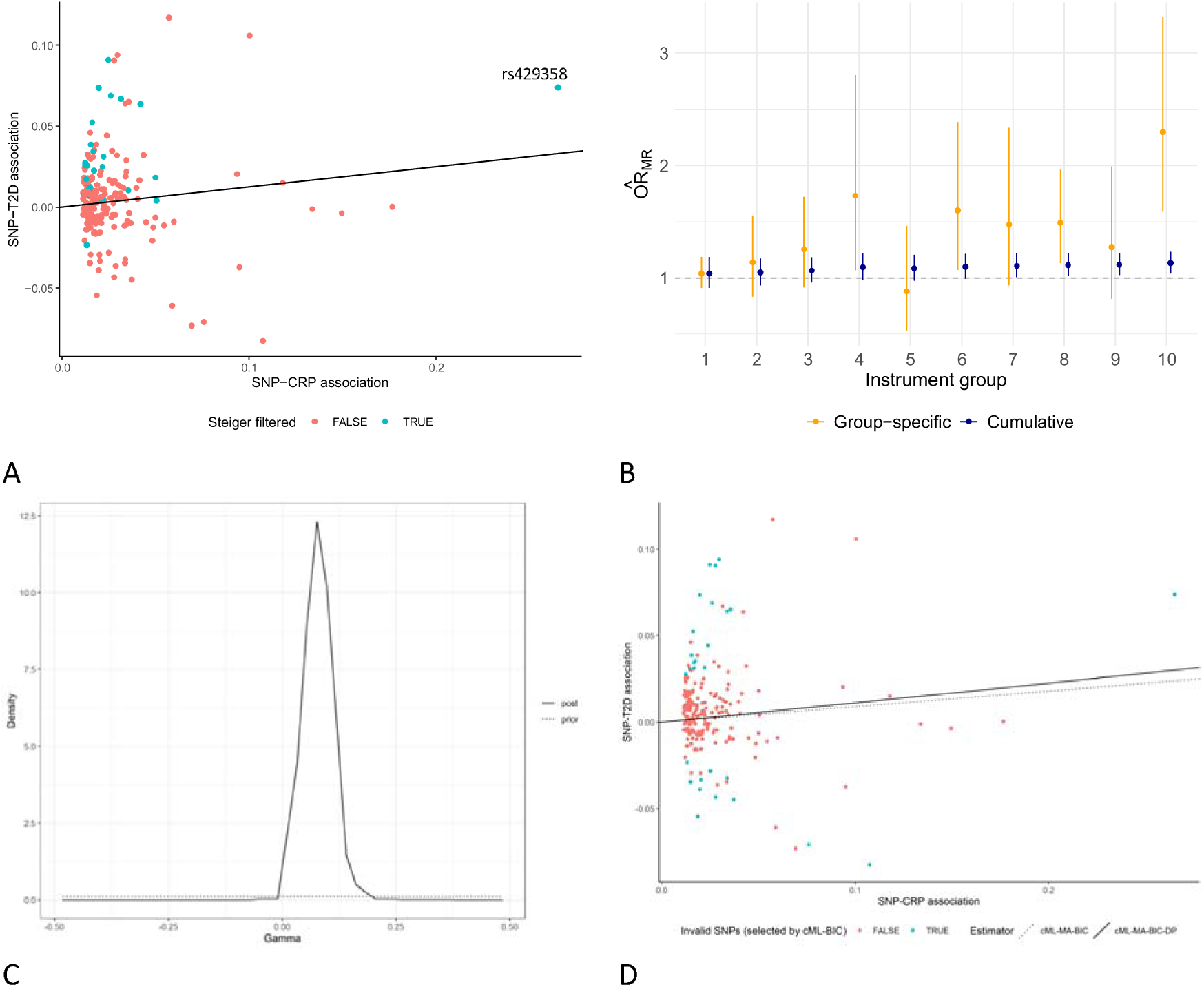
MR results from univariable MR estimation in estimation of the effect of CRP on Type 2 Diabetes. A. A Scatter plot of SNP-exposure and SNP-outcome associations (in log odds) for the SNPs used in the MR estimation of the effect of CRP on type 2 diabetes (T2D). All SNPs are genome-wide significantly associated with CRP, those SNPs that showed a stronger association with at least one of the top 10 potential confounders identified are highlighted in blue. The fitted line shows the IVW effect estimate using all SNPS. B. MR Corge per group IVW effect estimates (in odds ratios) for the effect of CRP on T2D. SNPs are grouped evenly by their association with CRP (18 or 19 SNPs per group). Group 1 has the strongest association with CRP. Yellow data points show the estimate and 95% confidence interval estimated using only the SNPs in that group, blue data points show the estimate and 95% confidence interval estimated using SNPs from that group and all groups that are more strongly associated with the exposure. Full results are given in Supplementary Table 7. C. Posterior and prior distribution of the causal effect estimate (gamma) in the MR CAUSE estimation of the effect of CRP on T2D. Full results are given in Supplementary Table 8. D. A Scatter plot of SNP-exposure and SNP-outcome associations (in log odds) for the SNPs used in the MR estimation of the effect of CRP on type 2 diabetes (T2D). SNPs identified as invalid in a cML-BIC estimation are highlighted in blue. The dotted fitted line shows the cML-MA-BIC estimate. The solid fitted line shows the cML-MA-BIC-DP estimate. Correlation of the GWAS summary statistics (ρ) was estimated using the intercept from a fitted bivariate LD-score regression model (LDSC) and accounted for in the cML estimation. Full results are given in Supplementary Table 9.

The protein traits identified in the top 10 confounders were all associated with the same variant (*rs429358* located in the APOE locus), raising the possibility that these associations could reflect either horizontal pleiotropy or, less likely, mediation through CRP. rs429358 has a substantial influence on CRP (highlighted in **Figure 5a**, beta: 0.265, p-value 1.0×10^-200^) and when it was included in the analysis (i.e. when the adiposity related confounders were considered in the Steiger filtering but not the protein markers) the weighted mode estimator association departed from the null.

This pattern was further supported by MR Corge analysis[27] (**Figure 5b**). MR Corge groups variants by the strength of their association (measured by the beta effect estimated from the GWAS study) with CRP and estimates MR effect estimates by group both individually and by combining groups cumulatively, starting with the strongest group. The group of variants most strongly associated with CRP (Group 1) showed no evidence of an effect on T2D. Larger, more precise effects only emerge as additional, weaker instruments are added. Notably, Group 1 included *rs2211320*, a variant within the CRP gene locus.

Recently developed methods Causal Analysis Using Summary Effect estimates (CAUSE)[14] and Mendelian Randomization with constrained Maximum Likelihood (MR-cML) [16, 41] aim to account for correlated pleiotropy in the estimates obtained. We explore whether these estimators can account for the likely presence of heritable confounding in our application (**Figures 5c and 5d, Supplementary Tables 8 and 9**). CAUSE applies a mixture model which allows for a minority of variants to be subject to correlated pleiotropy while the remainder may exhibit uncorrelated pleiotropy. Rather than focusing on genome-wide significant variants, CAUSE uses a wider set of LD-pruned variants across the genome. **Figure 5c** shows the estimated posterior distribution for the causal effect of CRP on T2D, with a median effect (in log odds) of 0.08 (95% CI: 0.01, 0.15). This is similar to estimates obtained with IVW, weighted Median and weighted Mode estimators without any Steiger filtering (**Table 3**), and thus indicates that CAUSE may not be able to account for heritable confounding in this application.

MR-cML is based on the assumption that the valid IVs are in the plurality, i.e. that they form the largest group among the sets of different Wald ratio estimates. It applies a constrained maximum likelihood estimator where the number of IVs invalid due to pleiotropy is constrained to a given integer which is selected using a Bayesian information criterion (BIC). The set of invalid variants suggested by this estimator is highlighted on a scatterplot in **Figure 5d**, with fitted lines illustrating the effects estimated by model averaging versions of MR-cML. Effect estimates (in log odds) range between 0.09 and 0.11 for different versions of the estimator. These are in line with the estimates obtained with CAUSE as well as traditional univariable MR estimators without Steiger filtering, suggesting that MR-cML is unable to address heritable confounding in the given application.

## Discussion

In this paper we have shown how correlated pleiotropy due to heritable confounding can bias MR effect estimates when the genetic variants selected as instruments are associated with a confounder of the exposure and outcome of interest. We show that the bias in MR from including instruments associated with a heritable confounder could be greater than the bias induced by that confounder in linear regression. Through theory and in our network simulations we predict that as GWAS sample sizes increase genetic variants identified as associated with an exposure are increasingly likely to be associated with an upstream confounder of the exposure and outcome. This will increase the presence of correlated pleiotropy and heritable confounding in MR effect estimates.

The strategy of adjusting for confounding in linear regression can be unreliable where incomplete coverage of confounders and imperfect measurement of those confounders are likely to lead to biased estimates. MR is often motivated as a method that sidesteps the need for such adjustment approaches.[3, 4, 42, 43] Yet here we argue that MR estimates are increasingly likely to be susceptible to bias due to confounding mechanisms, especially as larger GWAS sample sizes yield putative instruments that are more distal to the exposure, and that an adjustment strategy is required for MR also. There are three important ways in which adjustment in MR differ from adjustment in linear regression. First, because MR can be performed using GWAS summary statistics, it is possible to search a considerably greater phenotypic space for potential confounding paths than would be possible when restricting analysis to individual level data, as is required in linear regression.[44] Second, there is no requirement that putative confounders are perfectly measured, as estimation of the confounding effect through IV methods can mitigate such measurement error issues. Finally, MVMR is not subject to collider bias in the same way as linear regression when confounders are mis-specified, although misspecification of a mediator as a confounder can lead to incorrect interpretation of the results obtained.[25]

If a substantial proportion of the genetic instruments for an exposure are associated with a confounder of the exposure and outcome, correlated pleiotropy can bias all of the standard pleiotropy robust methods of estimation. In this case each pleiotropy robust method may be biased in the same way as the IVW estimator, potentially leading to false confidence in the results obtained. These results are likely to particularly apply to exposures that are downstream of a phenotype (such as BMI) which is associated with many genetic variants and has an influence on a wide range of traits, or where there are likely to be relatively few (if any) genetic variants more directly associated with the exposure of interest. The results given here highlight why it is important to consider whether the IV assumptions are likely to hold even when a range of MR estimates give consistent results for the effect of the exposure on the outcome.

We show that the bias from heritable confounding with a known confounder could be corrected through the application of either MVMR including the confounder, or Steiger filtering to remove any genetic variants that explain more variation in the confounder than the exposure. We therefore recommend that researchers applying MR with summary statistics generated from large GWAS studies check the association of their SNPs with the main potential confounders of the exposure and the outcome. This could be done either through a PheWAS approach as taken here or by considering a set of hypothesised potential confounders. Steiger filtering, removing SNPs that explain more variation in the confounder than the exposure, and/or MVMR also including the confounder can then be applied to the MR analysis as an additional robustness test. A substantial difference between the results obtained is an indication of bias in the original MR analysis. However, in practice it is likely that confounders will not be identifiable/measured and there will be a number of confounders acting in this way, so such an approach does not guarantee the removal of all bias from heritable confounding.

There are limitations to this approach as it requires identifying potential confounders and obtaining GWAS results for those confounders. Steiger filtering may not identify variants as being more strongly associated with a confounder if the GWAS for the confounder has substantially less power than the GWAS for the exposure or the variation explained by a variant in the exposure and confounder is very similar.[26] Steiger filtering relies on a estimate of the R^2^ on the liability scale for binary traits, adding a layer of uncertainty in the comparison when binary and continuous traits are being compared. However the limitations of Steiger filtering and potential biasing mechanisms have been explored at length elsewhere[26, 45] and its sensitivity to measurement error and complex confounding is generally low.

There are also limitations in the application of MVMR as weak instruments could lead to a change in the effect estimates in the absence of heritable confounding and therefore it is important to assess the robustness of the MVMR estimation when applying this method.[25, 40] These relative strengths and weaknesses were highlighted in our application; MVMR could not be applied to most of the protein biomarkers because of weak instruments and showed little attenuation of the effect in the one protein biomarker it was applied to (Tubulin-specific chaperone A). This is most likely because although the conditional F-statistics were larger than 10 Tubulin-specific chaperone A was still relatively weakly predicted with a conditional F-statistic of 11.18. showed a substantial attenuation of the effect when it was applied to adiposity related traits, suggesting that when the exposures are strongly predicted it can more fully adjust for the bias from heritable confounding. Another limitation in the application of MVMR is that caution needs to be applied when assessing potential heritable confounders and avoiding adjusting for mediators of the effect of the exposure on the outcome. When adjusting for a mediator the estimand targeted by MVMR differs from that targeted by MR and differences in that effect estimate obtained could be due to mediation rather than heritable confounding. All the traits considered as potential confounders in our application showed evidence of a causal effect on our exposure. Additional sensitivity analyses could be applied to ensure that any potential heritable confounders are not mediators (for example by estimating the effect of the exposure on the potential heritable confounder).

As with all MR methods the limitations of the approaches proposed here mean it is vital for researchers to consider the nature of the data being used, consider applying a range of methods, and where possible triangulate against other approaches to estimation.[6, 46] Other approaches that could be applied to help identify bias from heritable confounding include cis MR and methods that adjust for correlated pleiotropy. Cis MR makes use of genetic variants that are in a gene region known to affect a trait such as a protein.[47, 48] This concept can be extended to any setting where the exposure has a clear biological mechanism by which some genetic variants affect the exposure. Where possible comparing the results obtained for the MR estimation with all genetic variants to the results obtained for the genetic variants which have a known biological pathway can help to support the results or highlight the potential for bias in the MR estimate. More broadly, understanding the genetic variants being used as instruments can help to ensure that MR analyses are conducted robustly.[6] Recently novel MR methods have been proposed that attempt to adjust for correlated pleiotropy.[14, 16–19, 49] Each of these methods depends on different assumptions about the nature of the correlated pleiotropy and applies a different approach to correct for that pleiotropy. Applying these in addition to the approaches proposed here to identify specific confounders will help to identify whether results are biased by confounding. Approaches such as MRClust that cluster the effect estimates obtained by individual SNPs can also help identify groups of SNPs that are biased by the same heritable confounder if they identify a separate cluster for those SNPs.[50]

Heritable confounding leads to many issues with respect to gene-environment equivalence (G-EE). The G-EE assumption is that changing the exposure through an intervention on the genetic variants used as instruments would produce the same effect on the outcome as would manipulating the exposure through implementing certain interventions.[4, 5, 51, 52] When a variant influences the exposure through an upstream phenotype, then the hypothetical counterfactual change in the variant would produce the effects of the change in the upstream phenotype (which would include a change in the exposure of interest), rather than that of changing the exposure of interest alone. The CRP example suggests that cis CRP variants may mimic differences in circulating CRP levels produced by interventions such as antisense oligonucleotides to CRP or selective CRP apheresis.[53, 54] However variants which influence CRP through adiposity or a range of protein biomarkers will not produce an effect that is equivalent to the selective targeting of CRP levels. G-EE is simply implausible for many exposures that are currently being investigated using MR[5], as it is unlikely that there are any genetic variant influences on the exposure which are not acting through an upstream phenotype, and thus liable to produce heritable confounding. The caveat that genetic influences are from conception onwards, whereas interventions in general occur later in life, also applies when considering the reasonableness – or otherwise – of G-EE.[2, 4, 5, 43]

In our illustrative example we estimate the effect of CRP on T2D, systematically identifying potential confounders through a PheWAS approach. This application identified that the observed effect in the MR estimation may be driven by heritable confounding from adiposity, an established risk factor for elevated CRP levels.[55] Notably, a single genomic region tagged by rs429358 in the *APOE* locus emerged as a particularly influential locus in this analysis and was associated with many potential confounders. The APOE locus is a well established locus for Alzheimer’s disease[56] and has been associated with a range of different traits including lipid metabolism[57] and changes in BMI during adulthood[58]. Recent research has suggested that it may be a pleiotropic immune modulator[59] associated with many different diseases. This collective evidence strongly raises the possibility that it is a heritable confounder with the potential to bias our MR estimation. The prominence of this variant within the analysis underscores how increasing GWAS sample size, while increasing power to detect novel variants, also enables identification of genetic variants with more complex trait associations that are more likely to exhibit pleiotropic effects, increasing the risk of heritable confounding in MR analyses.

While our results show that the MR results can be more biased than linear regression from the exclusion of a single heritable confounder there are many other sources of bias for both types of estimation that should be considered when determining how much weight to give to the results from each type of analysis. It is likely that there will be a number of confounders of the exposure and outcome, if some of these are not associated with the SNPs used as instruments then the MR estimates may still be less biased from confounding than the observational linear regression estimates. Additionally, MR can overcome bias and effect attenuation from measurement error that occurs in linear regression for continuous exposures and can improve bias from mismeasured binary exposures.[60] However, MR is subject to other forms of bias such as population stratification and pleiotropy of other forms which should also be considered in any analysis.

A limitation of the simulation results here is how we have set up the structure of relationships between genetic variants and phenotypes. In our network simulations we assumed that every edge has the same strength of effect. In our simulations of the bias of the different MR estimators we have assumed the same distribution in size of the association between the relevant genetic variants and the exposure and confounder. These assumptions are relatively strong and are not likely to be realistic for every trait. For some traits the confounders will be affected more strongly by genetic variants than the exposure, which will accentuate the biases described here. For other traits the direct effect of the genetic variants on the exposure will be much larger than for the confounders, meaning that bias of the type described here will be less prevalent. Which of these is likely to hold, along with how strongly the confounders are likely to affect the outcome, will determine how much of a problem the results described here are likely to be in any particular case. Each MR study conducted should therefore consider individually how likely there is to be bias from heritable confounding in that setting and what the main confounders causing that bias may be. Feedback loops were not considered in our simulations and future work should examine how they manifest and potentially impact cross-sectional MR studies. Findings from MR studies should be interpreted within a triangulation of evidence framework, including falsification strategies such as the use of negative controls, which may trigger further investigation to identify the source of any observed inconsistencies.[61]

## Supporting information

Supplementary material section 1

Supplementary material section 2

## Acknowledgements

ES is funded by the Medical Research Council (UKRI077). ES, NV, TP, GH, GDS, KT work in a unit funded by the Medical Research Council (MC_UU_00032/1, MC_UU_00032/2). This research has been conducted using the UK Biobank Resource under Application Number 81499.

## Data availability

Summary statistics used in all applied analyses are available from the MRC IEU OpenGWAS platform (doi: 10.1101/2020.08.10.244293 and https://doi.org/10.7554/eLife.34408). Phenotypic correlations of CRP and T2D were estimated using data from the UK Biobank which are available to researchers upon application (https://www.ukbiobank.ac.uk/). UK Biobank has received ethical approval from the NHS North West Centre for Research Ethics Committees (references: 11/NW/0382, 16/NW/0274, 21/NW/0157).

## Code availability

The code for the simulations and analyses is available at https://github.com/eleanorsanderson/confounding.

## References

1. Davey Smith, G. and A.N. Phillips, Correlation without a cause: an epidemiological odyssey. International Journal of Epidemiology, 2020. 49(1): p. 4–14.

2. Davey Smith, G. and S. Ebrahim, ‘Mendelian randomization’: can genetic epidemiology contribute to understanding environmental determinants of disease?*. International Journal of Epidemiology, 2003. 32(1): p. 1–22.

3. Richmond, R. and G. Davey Smith, Mendelian randomization: concepts and scope. Cold Spring Harbour Perspectives in Medicine, 2021.

4. Sanderson, E., et al., Mendelian randomization. Nature Reviews Methods Primers, 2022. 2(1): p. 6.

5. Davey Smith, G., G. Hemani, and S. Ebrahim, Gene-environment equivalence: The fundamental principle of Mendelian randomization. PLOS Medicine, 2026. 23(3): p. e1005013.

6. Sanderson, E., et al., Challenges and future directions for Mendelian randomization. Nature Genetics, 2026: p. 1–11.

7. O’Connor, L.J., et al., Extreme Polygenicity of Complex Traits Is Explained by Negative Selection. The American Journal of Human Genetics, 2019. 105(3): p. 456–476.

8. Yengo, L., et al., A saturated map of common genetic variants associated with human height. Nature, 2022. 610(7933): p. 704–712.

9. Boyle, E.A., Y.I. Li, and J.K. Pritchard, An Expanded View of Complex Traits: From Polygenic to Omnigenic. Cell, 2017. 169(7): p. 1177–1186.

10. Nam, Y., et al., Harnessing artificial intelligence in multimodal omics data integration: paving the path for the next frontier in precision medicine. Annual review of biomedical data science, 2024. 7.

11. Davey Smith, G. and G. Hemani, Mendelian randomization: genetic anchors for causal inference in epidemiological studies. Hum Mol Genet, 2014. 23(R1): p. R89–98.

12. Brumpton, B., et al., Within-family studies for Mendelian randomization: avoiding dynastic, assortative mating, and population stratification biases. Nature Communications, 2020. 11(1): p. 1–13.

13. Davies, N.M., et al., Within family Mendelian randomization studies. Human Molecular Genetics, 2019. 28(R2): p. R170–R179.

14. Morrison, J., et al., Mendelian randomization accounting for correlated and uncorrelated pleiotropic effects using genome-wide summary statistics. Nature Genetics, 2020: p. 1–7.

15. Cheng, Q., et al., MR-LDP: a two-sample Mendelian randomization for GWAS summary statistics accounting for linkage disequilibrium and horizontal pleiotropy. NAR genomics and bioinformatics, 2020. 2(2): p. lqaa028–lqaa028.

16. Xue, H., X. Shen, and W. Pan, Constrained maximum likelihood-based Mendelian randomization robust to both correlated and uncorrelated pleiotropic effects. The American Journal of Human Genetics, 2021. 108(7): p. 1251–1269.

17. Xu, S., W.K. Fung, and Z. Liu, MRCIP: a robust Mendelian randomization method accounting for correlated and idiosyncratic pleiotropy. Briefings in Bioinformatics, 2021. 22(5).

18. Brown, B.C. and D.A. Knowles, Welch-weighted Egger regression reduces false positives due to correlated pleiotropy in Mendelian randomization. The American Journal of Human Genetics, 2021. 108(12): p. 2319–2335.

19. Cheng, Q., et al., Mendelian randomization accounting for complex correlated horizontal pleiotropy while elucidating shared genetic etiology. Nature Communications, 2022. 13(1): p. 6490.

20. Cheng, Q., et al., MR-Corr2: a two-sample Mendelian randomization method that accounts for correlated horizontal pleiotropy using correlated instrumental variants. Bioinformatics, 2022. 38(2): p. 303–310.

21. Lin, Z., Y. Deng, and W. Pan, Combining the strengths of inverse-variance weighting and Egger regression in Mendelian randomization using a mixture of regressions model. PLoS genetics, 2021. 17(11): p. e1009922.

22. Darrous, L., N. Mounier, and Z. Kutalik, Simultaneous estimation of bi-directional causal effects and heritable confounding from GWAS summary statistics. Nat Commun, 2021. 12(1): p. 7274.

23. Darrous, L., et al., PheWAS-based clustering of Mendelian Randomisation instruments reveals distinct mechanism-specific causal effects between obesity and educational attainment. Nat Commun, 2024. 15(1): p. 1420.

24. Wang, J., et al., Causal Inference for Heritable Phenotypic Risk Factors Using Heterogeneous Genetic Instruments. PLoS genetics, 2021. 17(6).

25. Sanderson, E., et al., An examination of multivariable Mendelian randomization in the single-sample and two-sample summary data settings. International journal of epidemiology, 2019. 48(3): p. 713–727.

26. Hemani, G., K. Tilling, and G.D. Smith, Orienting the causal relationship between imprecisely measured traits using GWAS summary data. PLoS genetics, 2017. 13(11): p. e1007081.

27. Zhang, W., et al., MR Corge: sensitivity analysis of Mendelian randomization based on the core gene hypothesis for polygenic exposures. Bioinformatics, 2024. 40(11).

28. Burgess, S., A. Butterworth, and S.G. Thompson, Mendelian Randomization Analysis With Multiple Genetic Variants Using Summarized Data. Genetic Epidemiology, 2013. 37(7): p. 658–665.

29. Bowden, J., G. Davey Smith, and S. Burgess, Mendelian randomization with invalid instruments: effect estimation and bias detection through Egger regression. International journal of epidemiology, 2015. 44(2): p. 512–525.

30. Bowden, J., et al., Consistent estimation in Mendelian randomization with some invalid instruments using a weighted median estimator. Genetic epidemiology, 2016. 40(4): p. 304–314.

31. Hartwig, F.P., G. Davey Smith, and J. Bowden, Robust inference in summary data Mendelian randomization via the zero modal pleiotropy assumption. International journal of epidemiology, 2017. 46(6): p. 1985–1998.

32. Burgess, S., F. Dudbridge, and S.G. Thompson, Re:“Multivariable Mendelian randomization: the use of pleiotropic genetic variants to estimate causal effects”. American journal of epidemiology, 2015. 181(4): p. 290–291.

33. Zhu, M., et al., C-reactive protein and cancer risk: a pan-cancer study of prospective cohort and Mendelian randomization analysis. BMC Medicine, 2022. 20(1): p. 301.

34. Zhou, A. and E. Hyppönen, Vitamin D deficiency and C-reactive protein: a bidirectional Mendelian randomization study. International Journal of Epidemiology, 2022. 52(1): p. 260–271.

35. Han, X., et al., Using Mendelian randomization to evaluate the causal relationship between serum C-reactive protein levels and age-related macular degeneration. European Journal of Epidemiology, 2020. 35(2): p. 139–146.

36. Timpson, N.J., et al., C-reactive protein and its role in metabolic syndrome: mendelian randomisation study. Lancet, 2005. 366(9501): p. 1954–9.

37. Said, S., et al., Genetic analysis of over half a million people characterises C-reactive protein loci. Nature Communications, 2022. 13(1): p. 2198.

38. Watanabe, K., et al., Functional mapping and annotation of genetic associations with FUMA. Nature Communications, 2017. 8(1): p. 1826.

39. Elsworth, B., et al., The MRC IEU OpenGWAS data infrastructure. bioRxiv, 2020: p. 2020.08.10.244293.

40. Sanderson, E., W. Spiller, and J. Bowden, Testing and correcting for weak and pleiotropic instruments in two-sample multivariable mendelian randomisation. Statistics in medicine, 2021.

41. Lin, Z., H. Xue, and W. Pan, Combining Mendelian randomization and network deconvolution for inference of causal networks with GWAS summary data. PLOS Genetics, 2023. 19(5): p. e1010762.

42. VanderWeele, T.J., et al., Methodological Challenges in Mendelian Randomization. Epidemiology, 2014. 25(3): p. 427–435.

43. Davey Smith, G. and S. Ebrahim, Mendelian randomization: prospects, potentials, and limitations. International journal of epidemiology, 2004. 33(1): p. 30–42.

44. Wang, Q., et al., A phenome-wide bidirectional Mendelian randomization analysis of atrial fibrillation. International journal of epidemiology, 2022. 51(4): p. 1153–1166.

45. Hemani, G., et al., Sensitivity analyses gain relevance by fixing parameters observable during the empirical analyses. Genet Epidemiol, 2023. 47(6): p. 461–462.

46. Lawlor, D.A., K. Tilling, and G. Davey Smith, Triangulation in aetiological epidemiology. International Journal of Epidemiology, 2017. 45(6): p. 1866–1886.

47. Schmidt, A.F., et al., Genetic drug target validation using Mendelian randomisation. Nature communications, 2020. 11(1): p. 3255–3255.

48. Holmes, M.V., et al., Integrating genomics with biomarkers and therapeutic targets to invigorate cardiovascular drug development. Nature Reviews Cardiology, 2021. 18(6): p. 435–453.

49. Qi, G. and N. Chatterjee, Mendelian randomization analysis using mixture models for robust and efficient estimation of causal effects. Nature Communications, 2019. 10(1): p. 1941.

50. Foley, C.N., et al., MR-Clust: clustering of genetic variants in Mendelian randomization with similar causal estimates. Bioinformatics, 2020. 37(4): p. 531–541.

51. Davey Smith, G., Epigenesis for epidemiologists: does evo-devo have implications for population health research and practice? International Journal of Epidemiology, 2012. 41(1): p. 236–247.

52. Skrivankova, V.W., et al., Strengthening the reporting of observational studies in epidemiology using mendelian randomisation (STROBE-MR): explanation and elaboration. bmj, 2021. 375.

53. Warren, M.S., et al., Results of a proof of concept, double-blind, randomized trial of a second generation antisense oligonucleotide targeting high-sensitivity C-reactive protein (hs-CRP) in rheumatoid arthritis. Arthritis Research & Therapy, 2015. 17(1): p. 80.

54. Ries, W., et al., C-Reactive Protein Apheresis as Anti-inflammatory Therapy in Acute Myocardial Infarction: Results of the CAMI-1 Study. Front Cardiovasc Med, 2021. 8: p. 591714.

55. Timpson, N.J., et al., C-reactive protein levels and body mass index: elucidating direction of causation through reciprocal Mendelian randomization. International Journal of Obesity, 2011. 35(2): p. 300–308.

56. Belloy, M.E., V. Napolioni, and M.D. Greicius, A quarter century of APOE and Alzheimer’s disease: progress to date and the path forward. Neuron, 2019. 101(5): p. 820–838.

57. Lumsden, A.L., et al., Apolipoprotein E (APOE) genotype-associated disease risks: a phenome-wide, registry-based, case-control study utilising the UK Biobank. eBioMedicine, 2020. 59: p. 102954.

58. Venkatesh, S.S., et al., Characterising the genetic architecture of changes in adiposity during adulthood using electronic health records. Nature Communications, 2024. 15(1): p. 5801.

59. Shvetcov, A., et al., APOE ε4 carriers share immune-related proteomic changes across neurodegenerative diseases. Nature Medicine, 2025. 31(8): p. 2590–2601.

60. Frazis, H. and M.A. Loewenstein, Estimating linear regressions with mismeasured, possibly endogenous, binary explanatory variables. Journal of Econometrics, 2003. 117(1): p. 151–178.

61. Gutierrez, S., M.M. Glymour, and G.D. Smith, Evidence triangulation in health research. Eur J Epidemiol, 2025. 40(7): p. 743–757.

62. Mbatchou, J., et al., Computationally efficient whole-genome regression for quantitative and binary traits. Nat Genet, 2021. 53(7): p. 1097–1103.

63. Yengo, L., et al., Meta-analysis of genome-wide association studies for height and body mass index in∼ 700000 individuals of European ancestry. Human molecular genetics, 2018. 27(20): p. 3641–3649.

