## Supplementary material section 1 for "Heritable confounding of exposure and outcome in Mendelian randomization studies"

1.1 **Relative bias from linear regression and IV:**

The model we consider can be written as;

$$Y=\beta_{0y}+ \beta_{xy}X+ \beta_{uy}U+v_{y}$$

$$X= \beta_{0x}+ \pi_{zx}Z+ \beta_{ux}U+v_{x}$$

$$U= \beta_{0u}+ \pi_{zu}Z+v_{u}$$

Where $v_{y}, v_{x}$and $v_{u}$ are all uncorrelated random error terms. This model is illustrated in **Figure 2A** in the main paper.

In this model the estimate of the effect of the exposure on the outcome from linear regression is given by;

$$\hat{\beta}_{LR}= \frac{\sum XY}{\sum X^{2}}$$

$$\hat{\beta}_{LR}= \frac{\sum X(\beta_{xy}X+ \beta_{uy}U+v_{y})}{\sum X^{2}}$$

$$\hat{\beta}_{LR}= \beta_{xy}+\frac{\sum{X(\beta}_{uy}U+v_{y})}{\sum X^{2}}$$

$$\hat{\beta}_{LR}= \beta_{xy}+\beta_{uy}\frac{\sum XU}{\sum X^{2}}+\frac{\sum Xv_{y}}{\sum X^{2}}$$

As we have assumed no other confounders in this simple model $v_{y}$ is independent of the exposure $X$and so $E\left( \sum Xv_{y} \right)=0$. We can additionally define $\delta_{ux}$ as the estimate that would be obtained from a regression of $X$on $U.$Note: this will not be the same as $\beta_{ux}$ due to the confounding of $U$and $X$by $Z$ but will estimate the correlation between $X$and $U$. Therefore;

$$\begin{aligned} E\left( \hat{\beta}_{LR} \right)= \beta_{xy}+ \beta_{uy}\delta_{ux}\#\left( 1 \right) \end{aligned}$$

This can alternatively be expressed as;

$$E\left( \hat{\beta}_{LR} \right)=\beta_{xy}+\frac{\beta_{uy}}{\beta_{ux}}R_{ux}^{2}$$

Where $R_{ux}^{2}$ is the variation in $X$ explained by $U$. i.e. the true causal effect of the exposure on the outcome plus a bias term that depends on the effect of the confounder on the outcome and the correlation between the confounder and the exposure.

The estimate of the effect of the exposure on the outcome from IV estimation is given by;

$$\beta_{IV}= \frac{\sum ZY}{\sum ZX}$$

$$\hat{\beta}_{IV}= \frac{\sum Z(\beta_{xy}X+ \beta_{uy}U+v_{y})}{\sum ZX}$$

$$\hat{\beta}_{IV}=\beta_{xy}+ \frac{\sum Z( \beta_{uy}U+v_{y})}{\sum ZX}$$

$$\hat{\beta}_{IV}=\beta_{xy}+ \beta_{uy}\frac{\sum ZU}{\sum ZX}+ \frac{\sum Zv_{y}}{\sum ZX}$$

$$\hat{\beta}_{IV}=\beta_{xy}+ \beta_{uy}\frac{\sum ZU}{\sum Z(\pi_{zx}Z+ \beta_{xu}U+v_{x})}+ \frac{\sum Zv_{y}}{\sum ZX}$$

$$\hat{\beta}_{IV}=\beta_{xy}+ \beta_{uy}\frac{\sum Z(\pi_{zu}Z+ v_{u})}{\sum Z(\pi_{zx}Z+ \beta_{xu}(\pi_{zu}Z+ v_{u})+v_{x})}+ \frac{\sum Zv_{y}}{\sum ZX}$$

$$\hat{\beta}_{IV}=\beta_{xy}+ \beta_{uy}\frac{\sum(\pi_{zu}Z^{2}+ {Zv}_{u})}{\sum(\pi_{zx}Z^{2}+ \beta_{xu}(\pi_{zu}Z^{2}+ {Zv}_{u})+{Zv}_{x})}+ \frac{\sum Zv_{y}}{\sum ZX}$$

The instrument $Z$is independent and exogenous so taking expectations of this expression, $E\left( \sum Zv_{y} \right)=0$, $E\left( \sum Zv_{x} \right)=0$ and $E\left( \sum Zv_{u} \right)=0$. Therefore;

$$E\left( \hat{\beta}_{IV} \right)=\beta_{xy}+ \beta_{uy}\frac{\sum\pi_{zu}Z^{2}}{\sum{(\pi}_{zx}Z^{2}+ \beta_{xu}\pi_{zu}Z^{2})}$$

$$\begin{aligned} E\left( \hat{\beta}_{IV} \right)=\beta_{xy}+ \beta_{uy}\frac{\pi_{zu}}{\pi_{zx}+ \beta_{xu}\pi_{zu}} \#\left( 2 \right) \end{aligned}$$

This is the true causal effect of the exposure on the outcome plus a bias term that depends on the ratio of the pleiotropic effect of the instrument on the outcome ($\beta_{uy}\pi_{zu}$) to the effect of the instrument on the exposure ($\pi_{zx}+ \beta_{ux}\pi_{zu}$). i.e. the larger the pleiotropic effect as a proportion of the effect of the instrument on the exposure the larger the bias of the IV estimator. The bias in the MR estimate from a single heritable confounder will be less than the bias of the linear regression estimator from the same confounder when;

$$\frac{\beta_{uy}\pi_{zu}}{\pi_{zx}+ \beta_{ux}\pi_{zu}}\boldsymbol{<}\beta_{uy}\delta_{ux}$$

### Network simulations

We use the randomDAG function in the R/dagitty package to simulate a DAG with an arbitrary number of nodes, and a fixed probability that any set of nodes is connected. Nodes with no parents in the simulated DAG are identified as genetic variants, and 500 trait pairs are selected at random, in each pair assigning one to be the hypothesised exposure and one to be the hypothesised outcome. We then identify the path length for every genetic variant for the exposure, and also determine whether they act via a confounder, or otherwise (i.e. directly or through other traits that mediate the association between the genetic variant and the exposure and have no paths to the outcome). The effect size of the genetic variant on the exposure is $\beta^{L}$ where $L$ is the path length from genotype to exposure, and for simplicity every edge in the DAG has the same effect $\beta=0.2$. These simulations were conducted for a set of parameters of graph size N=75, 100, 125, 150 and graph density based on probability of an edge occurring ranging from p = 0.01, 0.02, …, 0.1. Each parameter combination was repeated 10 times.

**Results**

A genetic variant inducing correlated pleiotropy must influence a trait that is a parent to both exposure and outcome. We hypothesised that such a genetic variant would generally be associated with more traits upstream of the exposure than valid instruments, and therefore would have relatively smaller effects on the exposure than valid instruments. The consequence of such a relationship would be that increasing GWAS sample sizes to identify more instruments of smaller effect sizes could lead to increasing bias in MR estimates.

To explore whether this is likely to be the case we performed simulations in which we consider that all hypothesised exposure-outcome relationships exist in a broader genotype-phenotype network. The network represents a directed acyclic graph where all nodes are traits except those that have no parents, which are genetic variants. The simulation proceeds by randomly choosing two traits, one as exposure and one as outcome, and then evaluating a) which variants are instruments for the exposure, and whether or not the instruments act through confounders in the graph, and b) the path length from instrument to exposure. The length of the edge between any pair of nodes has been fixed to be the same length for all edges.

**Figure 3** illustrates that in this model instruments that act via confounders tend to have smaller effects due to more traits lying on the path from the genetic variant to the exposure. For traits where our simulation is a realistic model of the underlying biology an implication of this result is that as sample sizes grow for GWAS, the rate of discovery of genetic instruments that are liable to act via confounders will grow. However, it is possible that this will not hold for all traits, for some traits it may be that the strongest effects are via confounders (e.g. because those confounders have very strong effects on the exposure) and the genetic variants with smaller effects will be the unconfounded ones. It is therefore important to consider if this model is likely to hold for each exposure trait considered.

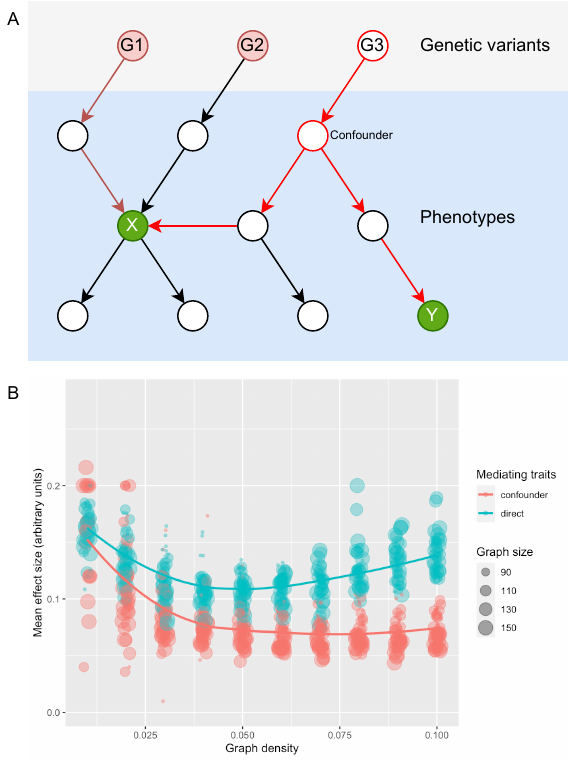

***Figure 3. Network-based simulations of the origin of instruments.***

A. *Illustration of a random network. For each possible trait pair we assign an exposure (X) and an outcome (Y). In this simplified example instrument G1 is considered to act ‘directly’ because there is no confounding path between X and Y, whereas G3 is considered to act via a confounding path. The effect size of each instrument is determined based on all edges having a uniform effect of 0.2.*
B. *The mean effect of instruments on the exposure (y-axis) across simulations (x-axis and point size), stratified by whether they act ‘directly’ or via confounding paths. We observe that in general direct instruments have larger effects than those acting via confounders.*
   1.2 **Details for Simulation to evaluate MR methods under heritable confounding**

### Simulations evaluating MR methods

We generated the data in this model for two separate samples. In each sample 400 SNPs were generated using a binomial distribution. The proportion of these SNPs associated with the mediator and the confounder varied across the simulations, varying between 50% associated with each trait to 12.5% associated with the mediator and 87.5% associated with the confounder. The SNP-trait (i.e. mediator/confounder) association was drawn from a normal distribution with mean 0 and standard deviation of 0.04 for each SNP, orientated so all SNPs had a positive association with the relevant trait.

The confounder was generated as the sum of all the confounder - SNP effects plus a random error term. The mediator was generated as the sum of all mediator – SNP effects plus a random error term. The exposure was generated as the sum of the mediator, the confounder and a random error term. As shown earlier, SNPs associated with a mediator may be more strongly associated with the exposure than SNPs associated with a confounder. Therefore, we down weighted the confounder – exposure association by 25% relative to the mediator – exposure association (by multiplying them by 0.75) to mimic this more distant association. Results with this down weighting varied (to 50% and 0%) are given in are given in **Supplementary Figure 1** and **Supplementary Figure 2** respectively . Finally, the outcome was generated as the effect of the confounder and a random error term. The expressions for the data generation process are given below.

In one of the samples generated we estimated the association between all of the SNPs and each of the exposure and confounder to generate GWAS summary statistics. This was repeated in the second sample for the SNPs and the outcome. We then select all SNPs (from either set) that were significantly associated with the exposure in the first sample at a defined p-value threshold to estimate the effect of the exposure on the outcome using summary-data MR methods. Following what is currently standard practice in summary-data MR the same sample was used for the SNP-exposure association in estimation as had been used to identify the SNPs associated with the exposure. We applied four common univariable MR methods of estimation: IVW[1], MR Egger[2], weighted median[3] and weighted mode[4]. We additionally estimated the model using multivariable MR (MVMR)[5, 6] including the confounder as an additional exposure, including all SNPs significantly associated with the confounder at the same p-value threshold. All univariable methods of estimation were repeated with Steiger filtering[7] applied between the exposure and confounder to remove SNPs that explain more variation in the confounder than the exposure.

Increasing the sample size of a GWAS will decrease the standard error in the SNP-trait association proportionally across all of the SNPs. This will lead to more SNPs being selected based on a fixed p-value threshold cut-off. Therefore, to model varying the sample size in the GWAS while keeping all other aspects of the data constant we varied the p-value threshold used to select SNPs for use as instruments in the MR across our simulations with larger p-values being equivalent to a larger sample size.

All models were simulated with a sample size of 100,000 for each of the exposure and outcome samples and 1000 repetitions.

A set of 400 SNPs were generated as;

$$G_{ij}\sim B(2, 0.4)$$

For individual ­$i$and SNP $j$. The SNP mediator/confounder effects were generated as;

$$\pi_{j}\sim N\left( 0, {0.04}^{2} \right)$$

The confounder was generated as;

$$C_{i}= \pi_{jc}G_{ij}+v_{c}$$

Where $v_{c}$ is a randomly normally distributed error term with mean 0 and standard deviation 1. $\pi_{jc}$ is an SNP effects vector for the confounder where the elements of $\pi_{j}$ associated with the mediator (rather than the confounder) have been set to 0.

The mediator was generated as;

$$M_{i}= \pi_{jm}G_{ij}+v_{c}$$

Where $v_{c}$ is a randomly normally distributed error term with mean 0 and standard deviation 1. $\pi_{jm}$ is an SNP effects vector for the mediator where the elements of $\pi_{j}$ associated with the confounder (rather than the mediator) have been set to 0.

The exposure was generated as;

$$X_{i}= M_{i}+0.75C_{i}+v_{x}$$

Where $v_{x}\sim N(0, 1)$.

The outcome was generated as;

$$Y_{i}=0.4C_{i}+v_{y}$$

Where $v_{y}\sim N(0, 1)$.

This data was generated for two different samples and summary statistics were generated by estimating the linear regressions;

$$C_{i}=\alpha_{c}+ {\beta_{cj}}G_{ij}+\epsilon_{c}$$

and

$$X_{i}= \alpha_{x}+\beta_{xj}G_{ij}+\epsilon_{x}$$

For all $j$ in the first sample and

$$Y_{i}=\alpha_{y}+ {\beta_{yj}}G_{ij}+\epsilon_{y}$$

For all $j$ in the second sample, to obtain estimates of $\beta_{cj}, \beta_{xj}$ and $\beta_{yj}$.

**Supplementary Figures**

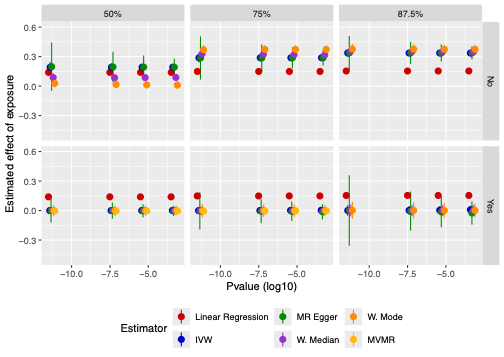

***Supplementary Figure 1 – Simulation of bias from a single heritable confounder in linear regression and MR when the confounder and mediator have equal effects on the exposure.***

*Results for the simulations varying; the proportion of the SNPs in G2 (50%, 75% or 87.5%), whether confounder adjustment (Steiger filtering/MVMR) is applied (No/Yes) and the p-value threshold applied (given on X-axis) to illustrate different GWAS sample sizes. Total number of SNPs – 400, % given is the % in G2, the rest are valid SNPs for X (i.e. G1). All SNPs that are significantly associated with X in the simulated data are included in the univariable MR estimation and all SNPs significantly associated with X or C are included in the MVMR estimation. There is no causal effect of X on Y, the effect of M on X is 1, of C on Y is 0.4 and of C on X is 0.75. n=10000, reps=1000. The effect of X on Y is estimated by; Linear regression, IVW, MR Egger, weighted median, weighted mode and MVMR including C as an additional exposure. Steiger filtered results includes all univariable estimators with Steiger filtering applied to remove SNPs that explain more variation in C than X.*

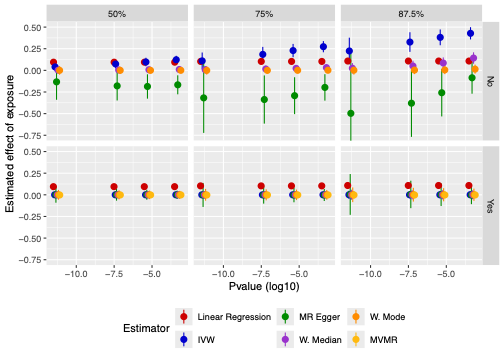

***Supplementary Figure 2 – Simulation of bias from a single heritable confounder in linear regression and MR when the confounder is more distal from the exposure than the mediator (effects downweighted by 50%).***

*Results for the simulations varying; the proportion of the SNPs in G2 (50%, 75% or 87.5%), whether confounder adjustment (Steiger filtering/MVMR) is applied (No/Yes) and the p-value threshold applied (given on X-axis) to illustrate different GWAS sample sizes. Total number of SNPs – 400, % given is the % in G2, the rest are valid SNPs for X (i.e. G1). All SNPs that are significantly associated with X in the simulated data are included in the univariable MR estimation and all SNPs significantly associated with X or C are included in the MVMR estimation. There is no causal effect of X on Y, the effect of M on X is 1, of C on Y is 0.4 and of C on X is 0.75. n=10000, reps=1000. The effect of X on Y is estimated by; Linear regression, IVW, MR Egger, weighted median, weighted mode and MVMR including C as an additional exposure. Steiger filtered results includes all univariable estimators with Steiger filtering applied to remove SNPs that explain more variation in C than X.*

| 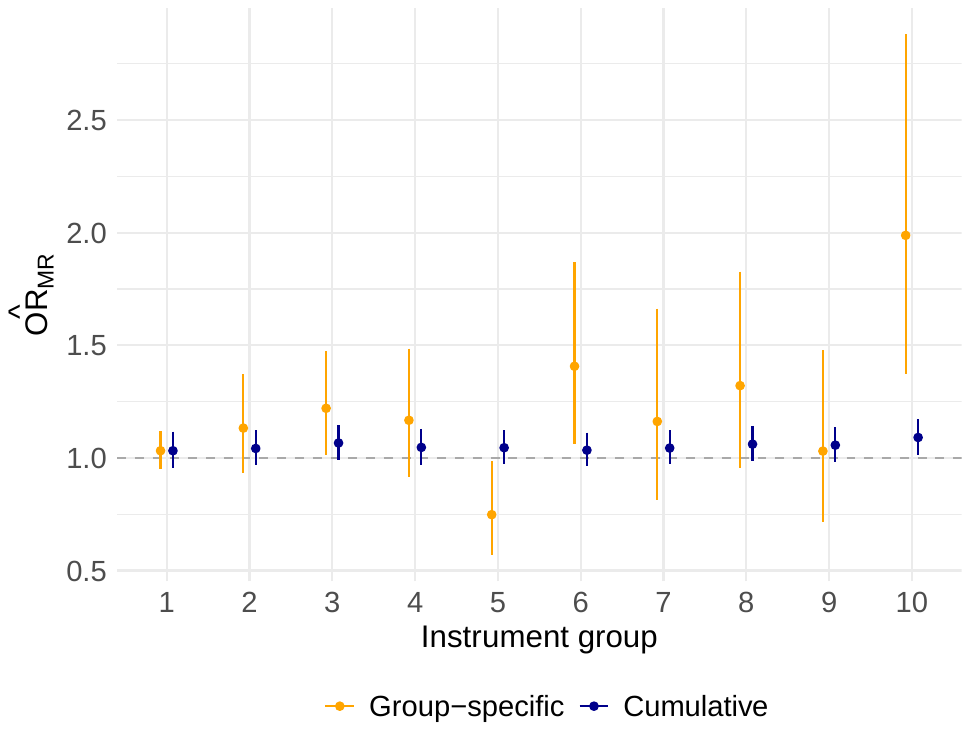 | 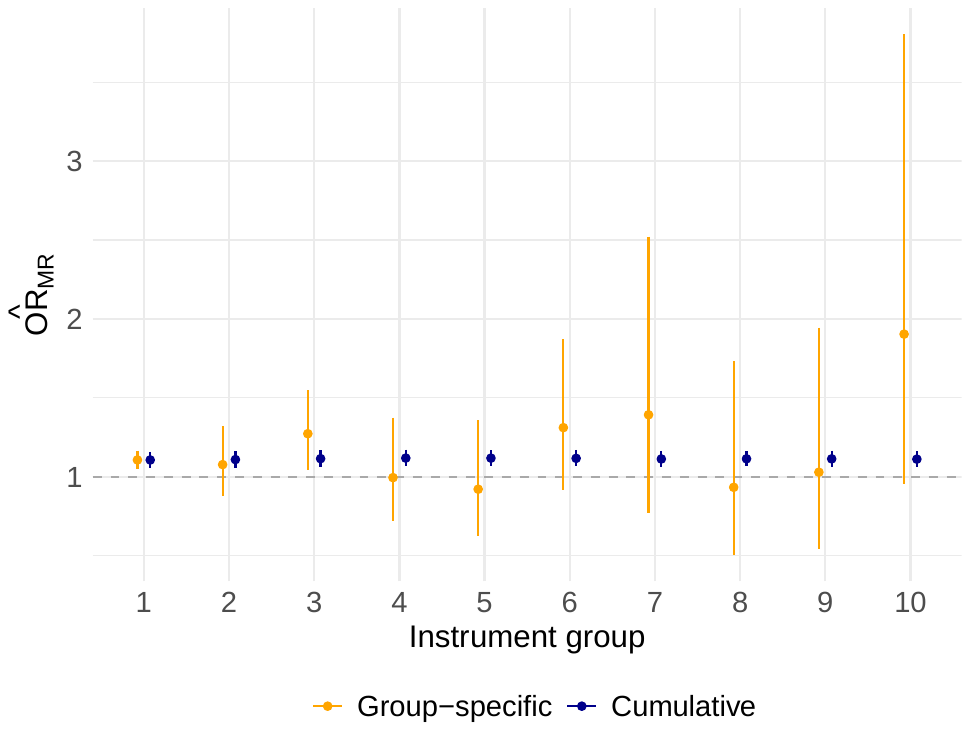 |
| --- | --- |
| (a) *Weighted median* | (b) *Weighted mode* |
| 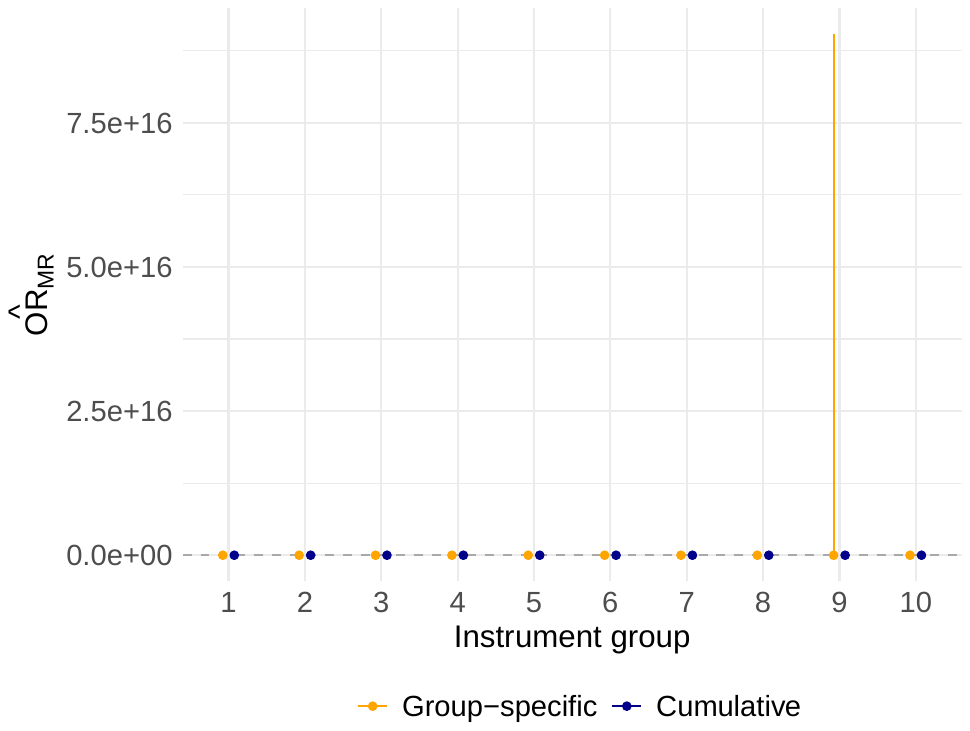 |  |
| (c) *MR Egger* |  |

***Supplementary Figure 4 – MR Corge plots for weighted median, weighted mode and MR Egger estimators for estimation of the effect of C Reactive protein on risk of Type 2 Diabetes.***

**Supplementary Tables**

***Supplementary Table 1 – Simulation results for univariable and multivariable MR including all SNPs selected as significantly associated with the exposure.***

|  | *Linear Regression* | | *IVW* | | *MR Egger* | | *Weighted Median* | | *Weighted Mode* | | *MVMR* | |
| --- | --- | --- | --- | --- | --- | --- | --- | --- | --- | --- | --- | --- |
| P-value | ***β*** | ***Std. Err*** | ***β*** | ***Std. Err*** | ***β*** | ***Std. Err*** | ***β*** | ***Std. Err*** | ***β*** | ***Std. Err*** | ***β*** | ***Std. Err*** |
| *50% of SNPs associated with confounder* | | | | |  |  |  |  |  |  |  |  |
| 5.00E-12 | 0.12 | 0.001 | 0.14 | 0.03 | -0.10 | 0.15 | 0.02 | 0.02 | 0.00 | 0.02 | 0.00 | 0.01 |
| 5.00E-08 | 0.12 | 0.001 | 0.16 | 0.03 | -0.05 | 0.09 | 0.03 | 0.01 | 0.00 | 0.01 | 0.00 | 0.01 |
| 5.00E-06 | 0.12 | 0.001 | 0.17 | 0.02 | -0.02 | 0.07 | 0.03 | 0.01 | 0.00 | 0.01 | 0.00 | 0.01 |
| 5.00E-04 | 0.12 | 0.001 | 0.18 | 0.02 | 0.02 | 0.05 | 0.03 | 0.01 | 0.00 | 0.01 | 0.00 | 0.01 |
| *75% of SNPs associated with confounder* | | | | |  |  |  |  |  |  |  |  |
| 5.00E-12 | 0.13 | 0.001 | 0.26 | 0.04 | -0.07 | 0.19 | 0.10 | 0.03 | 0.03 | 0.02 | 0.00 | 0.02 |
| 5.00E-08 | 0.13 | 0.001 | 0.29 | 0.03 | 0.03 | 0.11 | 0.15 | 0.03 | 0.05 | 0.03 | 0.00 | 0.01 |
| 5.00E-06 | 0.13 | 0.001 | 0.30 | 0.02 | 0.08 | 0.08 | 0.18 | 0.03 | 0.06 | 0.03 | 0.00 | 0.01 |
| 5.00E-04 | 0.13 | 0.001 | 0.31 | 0.02 | 0.14 | 0.06 | 0.21 | 0.02 | 0.08 | 0.03 | 0.00 | 0.01 |
| *87.5% of SNPs associated with confounder* | | | | |  |  |  |  |  |  |  |  |
| 5.00E-12 | 0.14 | 0.001 | 0.36 | 0.04 | 0.10 | 0.20 | 0.39 | 0.03 | 0.39 | 0.04 | 0.00 | 0.02 |
| 5.00E-08 | 0.14 | 0.001 | 0.38 | 0.03 | 0.19 | 0.11 | 0.41 | 0.02 | 0.45 | 0.04 | 0.00 | 0.02 |
| 5.00E-06 | 0.14 | 0.001 | 0.39 | 0.02 | 0.23 | 0.08 | 0.41 | 0.02 | 0.47 | 0.04 | 0.00 | 0.02 |
| 5.00E-04 | 0.14 | 0.001 | 0.40 | 0.02 | 0.28 | 0.06 | 0.41 | 0.02 | 0.47 | 0.03 | 0.00 | 0.02 |

*Results from the simulation of the model given in Figures 4, varying the proportion of SNPs associated with the exposure and confounder and the p-value used to select instruments. A larger P-value is equivalent to a larger sample size in the SNP discovery GWAS. IVW, MR Egger, Weighted median and Weighted mode are applied using all SNPs associated with the exposure at the p-value specified, MVMR estimation additionally includes all SNPs associated with the confounder. Results reported are the mean effect estimate and standard error across the repetitions. N=100,000, repetitions = 1000.*

***Supplementary Table 2 – Simulation results for univariable MR including all SNPs selected as significantly associated with the exposure with Steiger filtering applied to remove SNPs more strongly associated with the confounder than the exposure.***

|  | *Linear Regression* | | *IVW* | | *MR Egger* | | *Weighted Median* | | *Weighted Mode* | |
| --- | --- | --- | --- | --- | --- | --- | --- | --- | --- | --- |
| P-value | ***β*** | ***Std. Err*** | ***β*** | ***Std. Err*** | ***β*** | ***Std. Err*** | ***β*** | ***Std. Err*** | ***β*** | ***Std. Err*** |
| *50% of SNPs associated with confounder* | | | | |  |  |  |  |  |  |
| 5.00E-12 | 0.12 | 0.001 | 0.00 | 0.01 | 0.00 | 0.05 | 0.00 | 0.02 | 0.00 | 0.03 |
| 5.00E-08 | 0.12 | 0.001 | 0.00 | 0.01 | 0.00 | 0.04 | 0.00 | 0.01 | 0.00 | 0.03 |
| 5.00E-06 | 0.12 | 0.001 | 0.00 | 0.01 | 0.00 | 0.03 | 0.00 | 0.01 | 0.00 | 0.03 |
| 5.00E-04 | 0.12 | 0.001 | 0.00 | 0.01 | 0.00 | 0.03 | 0.00 | 0.01 | 0.00 | 0.02 |
| *75% of SNPs associated with confounder* | | | | |  |  |  |  |  |  |
| 5.00E-12 | 0.13 | 0.001 | 0.00 | 0.02 | 0.00 | 0.08 | 0.00 | 0.02 | 0.00 | 0.04 |
| 5.00E-08 | 0.13 | 0.001 | 0.00 | 0.01 | 0.00 | 0.06 | 0.00 | 0.02 | 0.00 | 0.03 |
| 5.00E-06 | 0.13 | 0.001 | 0.00 | 0.01 | 0.00 | 0.05 | 0.00 | 0.02 | 0.00 | 0.03 |
| 5.00E-04 | 0.13 | 0.001 | 0.00 | 0.01 | -0.01 | 0.04 | 0.00 | 0.02 | 0.00 | 0.03 |
| *87.5% of SNPs associated with confounder* | | | | |  |  |  |  |  |  |
| 5.00E-12 | 0.14 | 0.001 | 0.00 | 0.02 | 0.00 | 0.13 | 0.00 | 0.03 | 0.00 | 0.04 |
| 5.00E-08 | 0.14 | 0.001 | 0.00 | 0.02 | 0.00 | 0.09 | 0.00 | 0.03 | 0.00 | 0.04 |
| 5.00E-06 | 0.14 | 0.001 | 0.00 | 0.02 | 0.00 | 0.07 | 0.00 | 0.03 | 0.00 | 0.04 |
| 5.00E-04 | 0.14 | 0.001 | 0.00 | 0.02 | -0.01 | 0.06 | 0.00 | 0.03 | 0.00 | 0.04 |

*Results from the simulation of the model given in Figures 4, varying the proportion of SNPs associated with the exposure and confounder and the p-value used to select instruments. A larger P-value is equivalent to a larger sample size in the SNP discovery GWAS. IVW, MR Egger, Weighted median and Weighted mode are applied using all SNPs associated with the exposure at the p-value specified, MVMR estimation additionally includes all SNPs associated with the confounder. All MR estimation Steiger filtered to remove SNPs that explain more variation in the confounder than the exposure. Results reported are the mean effect estimate and standard error across the repetitions. N=100,000, repetitions = 1000.*

***Supplementary Table 3 – Simulation results for univariable and multivariable MR including all SNPs selected as significantly associated with the exposure. Mediator and confounder have the same size effect on the exposure.***

|  | *Linear Regression* | | *IVW* | | *MR Egger* | | *Weighted Median* | | *Weighted Mode* | | *MVMR* | |
| --- | --- | --- | --- | --- | --- | --- | --- | --- | --- | --- | --- | --- |
| P-value | ***β*** | ***Std. Err*** | ***β*** | ***Std. Err*** | ***β*** | ***Std. Err*** | ***β*** | ***Std. Err*** | ***β*** | ***Std. Err*** | ***β*** | ***Std. Err*** |
| *50% of SNPs associated with confounder* | | | | |  |  |  |  |  |  |  |  |
| 5.00E-12 | 0.14 | 0.001 | 0.19 | 0.03 | 0.20 | 0.13 | 0.09 | 0.02 | 0.03 | 0.02 | 0.00 | 0.01 |
| 5.00E-08 | 0.14 | 0.001 | 0.19 | 0.02 | 0.19 | 0.08 | 0.09 | 0.02 | 0.02 | 0.02 | 0.00 | 0.01 |
| 5.00E-06 | 0.14 | 0.001 | 0.19 | 0.02 | 0.19 | 0.06 | 0.09 | 0.02 | 0.01 | 0.02 | 0.00 | 0.01 |
| 5.00E-04 | 0.14 | 0.001 | 0.19 | 0.02 | 0.19 | 0.04 | 0.09 | 0.02 | 0.01 | 0.02 | 0.00 | 0.01 |
| *75% of SNPs associated with confounder* | | | | |  |  |  |  |  |  |  |  |
| 5.00E-12 | 0.15 | 0.001 | 0.29 | 0.02 | 0.29 | 0.11 | 0.33 | 0.02 | 0.37 | 0.02 | -0.01 | 0.02 |
| 5.00E-08 | 0.15 | 0.001 | 0.29 | 0.02 | 0.29 | 0.07 | 0.32 | 0.02 | 0.37 | 0.02 | 0.00 | 0.02 |
| 5.00E-06 | 0.15 | 0.001 | 0.29 | 0.02 | 0.29 | 0.05 | 0.32 | 0.01 | 0.37 | 0.02 | 0.00 | 0.01 |
| 5.00E-04 | 0.15 | 0.001 | 0.29 | 0.01 | 0.29 | 0.04 | 0.31 | 0.01 | 0.37 | 0.02 | 0.00 | 0.01 |
| *87.5% of SNPs associated with confounder* | | | | |  |  |  |  |  |  |  |  |
| 5.00E-12 | 0.15 | 0.001 | 0.34 | 0.02 | 0.34 | 0.09 | 0.36 | 0.02 | 0.38 | 0.03 | 0.00 | 0.02 |
| 5.00E-08 | 0.15 | 0.001 | 0.34 | 0.01 | 0.34 | 0.06 | 0.35 | 0.01 | 0.38 | 0.02 | 0.00 | 0.02 |
| 5.00E-06 | 0.15 | 0.001 | 0.34 | 0.01 | 0.34 | 0.04 | 0.35 | 0.01 | 0.37 | 0.02 | 0.00 | 0.02 |
| 5.00E-04 | 0.15 | 0.001 | 0.34 | 0.01 | 0.34 | 0.03 | 0.35 | 0.01 | 0.37 | 0.02 | 0.00 | 0.02 |

*Results from the simulation with equal effects of the confounder and mediator on the exposure, varying the proportion of SNPs associated with the exposure and confounder and the p-value used to select instruments. A larger P-value is equivalent to a larger sample size in the SNP discovery GWAS. IVW, MR Egger, Weighted median and Weighted mode are applied using all SNPs associated with the exposure at the p-value specified, MVMR estimation additionally includes all SNPs associated with the confounder. Results reported are the mean effect estimate and standard error across the repetitions. N=100,000, repetitions = 1000.*

***Supplementary Table 4 – Simulation results for univariable MR including all SNPs selected as significantly associated with the exposure with Steiger filtering applied to remove SNPs more strongly associated with the confounder than the exposure. Mediator and confounder have the same size effect on the exposure.***

|  | *Linear Regression* | | *IVW* | | *MR Egger* | | *Weighted Median* | | *Weighted Mode* | |
| --- | --- | --- | --- | --- | --- | --- | --- | --- | --- | --- |
| P-value | ***β*** | ***Std. Err*** | ***β*** | ***Std. Err*** | ***β*** | ***Std. Err*** | ***β*** | ***Std. Err*** | ***β*** | ***Std. Err*** |
| *50% of SNPs associated with confounder* | | | | |  |  |  |  |  |  |
| 5.00E-12 | 0.14 | 0.001 | 0.00 | 0.01 | 0.00 | 0.06 | 0.00 | 0.02 | 0.00 | 0.03 |
| 5.00E-08 | 0.14 | 0.001 | 0.00 | 0.01 | 0.00 | 0.04 | 0.00 | 0.01 | 0.00 | 0.03 |
| 5.00E-06 | 0.14 | 0.001 | 0.00 | 0.01 | 0.00 | 0.03 | 0.00 | 0.01 | 0.00 | 0.03 |
| 5.00E-04 | 0.14 | 0.001 | 0.00 | 0.01 | 0.00 | 0.03 | 0.00 | 0.01 | 0.00 | 0.02 |
| *75% of SNPs associated with confounder* | | | | |  |  |  |  |  |  |
| 5.00E-12 | 0.15 | 0.001 | 0.00 | 0.02 | 0.00 | 0.10 | 0.00 | 0.02 | 0.00 | 0.04 |
| 5.00E-08 | 0.15 | 0.001 | 0.00 | 0.02 | -0.01 | 0.06 | 0.00 | 0.02 | 0.00 | 0.03 |
| 5.00E-06 | 0.15 | 0.001 | 0.00 | 0.01 | -0.01 | 0.05 | 0.00 | 0.02 | 0.00 | 0.03 |
| 5.00E-04 | 0.15 | 0.001 | 0.00 | 0.01 | -0.01 | 0.04 | 0.00 | 0.02 | 0.00 | 0.03 |
| *87.5% of SNPs associated with confounder* | | | | |  |  |  |  |  |  |
| 5.00E-12 | 0.15 | 0.001 | 0.00 | 0.03 | 0.00 | 0.18 | 0.00 | 0.03 | 0.00 | 0.04 |
| 5.00E-08 | 0.15 | 0.001 | 0.00 | 0.02 | -0.01 | 0.10 | 0.00 | 0.03 | 0.00 | 0.04 |
| 5.00E-06 | 0.15 | 0.001 | 0.00 | 0.02 | -0.01 | 0.08 | 0.00 | 0.03 | 0.00 | 0.04 |
| 5.00E-04 | 0.15 | 0.001 | 0.01 | 0.02 | -0.03 | 0.06 | 0.00 | 0.03 | 0.00 | 0.04 |

*Results from the simulation with equal effects of the confounder and mediator on the exposure, varying the proportion of SNPs associated with the exposure and confounder and the p-value used to select instruments. A larger P-value is equivalent to a larger sample size in the SNP discovery GWAS. IVW, MR Egger, Weighted median and Weighted mode are applied using all SNPs associated with the exposure at the p-value specified. All MR estimation Steiger filtered to remove SNPs that explain more variation in the confounder than the exposure. Results reported are the mean effect estimate and standard error across the repetitions. N=100,000, repetitions = 1000.*

***Supplementary Table 5 – Simulation results for univariable and multivariable MR including all SNPs selected as significantly associated with the exposure. Confounder effect on exposure down weighted by 50%.***

|  | *Linear Regression* | | *IVW* | | *MR Egger* | | *Weighted Median* | | *Weighted Mode* | | *MVMR* | |
| --- | --- | --- | --- | --- | --- | --- | --- | --- | --- | --- | --- | --- |
| P-value | ***β*** | ***Std. Err*** | ***β*** | ***Std. Err*** | ***β*** | ***Std. Err*** | ***β*** | ***Std. Err*** | ***β*** | ***Std. Err*** | ***β*** | ***Std. Err*** |
| *50% of SNPs associated with confounder* | | | | |  |  |  |  |  |  |  |  |
| 5.00E-12 | 0.09 | 0.002 | 0.04 | 0.03 | -0.13 | 0.11 | 0.00 | 0.02 | 0.00 | 0.03 | 0.00 | 0.01 |
| 5.00E-08 | 0.09 | 0.002 | 0.07 | 0.03 | -0.18 | 0.09 | 0.01 | 0.01 | 0.00 | 0.02 | 0.00 | 0.01 |
| 5.00E-06 | 0.09 | 0.002 | 0.10 | 0.03 | -0.19 | 0.07 | 0.01 | 0.01 | 0.00 | 0.02 | 0.00 | 0.01 |
| 5.00E-04 | 0.09 | 0.002 | 0.12 | 0.02 | -0.17 | 0.06 | 0.01 | 0.01 | 0.00 | 0.01 | 0.00 | 0.01 |
| *75% of SNPs associated with confounder* | | | | |  |  |  |  |  |  |  |  |
| 5.00E-12 | 0.10 | 0.002 | 0.11 | 0.05 | -0.32 | 0.21 | 0.01 | 0.02 | 0.00 | 0.02 | 0.00 | 0.02 |
| 5.00E-08 | 0.10 | 0.002 | 0.18 | 0.04 | -0.34 | 0.14 | 0.02 | 0.02 | 0.00 | 0.02 | 0.00 | 0.01 |
| 5.00E-06 | 0.10 | 0.002 | 0.23 | 0.04 | -0.29 | 0.11 | 0.02 | 0.02 | 0.00 | 0.02 | 0.00 | 0.01 |
| 5.00E-04 | 0.10 | 0.002 | 0.27 | 0.03 | -0.20 | 0.08 | 0.03 | 0.02 | 0.00 | 0.02 | 0.00 | 0.01 |
| *87.5% of SNPs associated with confounder* | | | | |  |  |  |  |  |  |  |  |
| 5.00E-12 | 0.11 | 0.002 | 0.22 | 0.08 | -0.50 | 0.35 | 0.02 | 0.03 | 0.00 | 0.03 | 0.00 | 0.02 |
| 5.00E-08 | 0.11 | 0.002 | 0.33 | 0.06 | -0.38 | 0.20 | 0.05 | 0.03 | 0.00 | 0.02 | 0.00 | 0.02 |
| 5.00E-06 | 0.11 | 0.002 | 0.38 | 0.05 | -0.26 | 0.14 | 0.09 | 0.03 | 0.01 | 0.02 | 0.00 | 0.02 |
| 5.00E-04 | 0.11 | 0.002 | 0.43 | 0.04 | -0.09 | 0.09 | 0.14 | 0.03 | 0.01 | 0.03 | 0.00 | 0.02 |

*Results from the simulation with the effect of the confounder on the exposure down-weighted by 50% relative to the effect of the mediator on the exposure, varying the proportion of SNPs associated with the exposure and confounder and the p-value used to select instruments. A larger P-value is equivalent to a larger sample size in the SNP discovery GWAS. IVW, MR Egger, Weighted median and Weighted mode are applied using all SNPs associated with the exposure at the p-value specified, MVMR estimation additionally includes all SNPs associated with the confounder. Results reported are the mean effect estimate and standard error across the repetitions. N=100,000, repetitions = 1000.*

***Supplementary Table 6 – Simulation results for univariable MR including all SNPs selected as significantly associated with the exposure with Steiger filtering applied to remove SNPs more strongly associated with the confounder than the exposure. Confounder effect on exposure down weighted by 50%.***

|  | *Linear Regression* | | *IVW* | | *MR Egger* | | *Weighted Median* | | *Weighted Mode* | |
| --- | --- | --- | --- | --- | --- | --- | --- | --- | --- | --- |
| P-value | ***β*** | ***Std. Err*** | ***β*** | ***Std. Err*** | ***β*** | ***Std. Err*** | ***β*** | ***Std. Err*** | ***β*** | ***Std. Err*** |
| *50% of SNPs associated with confounder* | | | | |  |  |  |  |  |  |
| 5.00E-12 | 0.09 | 0.002 | 0.00 | 0.01 | 0.00 | 0.05 | 0.00 | 0.02 | 0.00 | 0.03 |
| 5.00E-08 | 0.09 | 0.002 | 0.00 | 0.01 | 0.00 | 0.03 | 0.00 | 0.01 | 0.00 | 0.03 |
| 5.00E-06 | 0.09 | 0.002 | 0.00 | 0.01 | 0.00 | 0.03 | 0.00 | 0.01 | 0.00 | 0.03 |
| 5.00E-04 | 0.09 | 0.002 | 0.00 | 0.01 | 0.00 | 0.02 | 0.00 | 0.01 | 0.00 | 0.02 |
| *75% of SNPs associated with confounder* | | | | |  |  |  |  |  |  |
| 5.00E-12 | 0.10 | 0.002 | 0.00 | 0.02 | 0.00 | 0.07 | 0.00 | 0.02 | 0.00 | 0.03 |
| 5.00E-08 | 0.10 | 0.002 | 0.00 | 0.01 | 0.00 | 0.05 | 0.00 | 0.02 | 0.00 | 0.03 |
| 5.00E-06 | 0.10 | 0.002 | 0.00 | 0.01 | 0.00 | 0.04 | 0.00 | 0.02 | 0.00 | 0.03 |
| 5.00E-04 | 0.10 | 0.002 | 0.00 | 0.01 | 0.00 | 0.04 | 0.00 | 0.02 | 0.00 | 0.03 |
| *87.5% of SNPs associated with confounder* | | | | |  |  |  |  |  |  |
| 5.00E-12 | 0.11 | 0.002 | 0.00 | 0.02 | 0.00 | 0.12 | 0.00 | 0.03 | 0.00 | 0.04 |
| 5.00E-08 | 0.11 | 0.002 | 0.00 | 0.02 | 0.00 | 0.08 | 0.00 | 0.03 | 0.00 | 0.04 |
| 5.00E-06 | 0.11 | 0.002 | 0.00 | 0.02 | 0.00 | 0.07 | 0.00 | 0.03 | 0.00 | 0.04 |
| 5.00E-04 | 0.11 | 0.002 | 0.00 | 0.02 | 0.00 | 0.05 | 0.00 | 0.03 | 0.00 | 0.04 |

*Results from the simulation with the effect of the confounder on the exposure down-weighted by 50% relative to the effect of the mediator on the exposure, varying the proportion of SNPs associated with the exposure and confounder and the p-value used to select instruments. A larger P-value is equivalent to a larger sample size in the SNP discovery GWAS. IVW, MR Egger, Weighted median and Weighted mode are applied using all SNPs associated with the exposure at the p-value specified. All MR estimation Steiger filtered to remove SNPs that explain more variation in the confounder than the exposure. Results reported are the mean effect estimate and standard error across the repetitions. N=100,000, repetitions = 1000.*

***Supplementary Table 7 – MR Corge results for estimation of the effect of CRP on Type 2 Diabetes***

|  |  |  | IVW | | | | Weighted Median | | | | Weighted mode | | | | MR Egger | | | |
| --- | --- | --- | --- | --- | --- | --- | --- | --- | --- | --- | --- | --- | --- | --- | --- | --- | --- | --- |
| group | N. snp | Fstat | beta | Std.Err. | 95% CI | | beta | Std. Err. | 95% CI | | beta | Std. Err. | 95% CI | | beta | Std. Err. | 95% CI | |
| Group Specific estimates | | |  |  |  |  |  |  |  |  |  |  |  |  |  |  |  |  |
| 1 | 23 | 1456.67 | 0.04 | 0.07 | (-0.10 | 0.17) | 0.03 | 0.04 | (-0.05 | 0.11) | 0.10 | 0.02 | (0.06 | 0.06) | 0.33 | 0.12 | (0.10 | 0.56) |
| 2 | 22 | 137.27 | 0.13 | 0.16 | (-0.18 | 0.44) | 0.12 | 0.10 | (-0.07 | 0.32) | 0.07 | 0.11 | (-0.15 | -0.15) | -1.25 | 2.38 | (-5.91 | 3.42) |
| 3 | 22 | 118.16 | 0.23 | 0.16 | (-0.09 | 0.54) | 0.20 | 0.09 | (0.02 | 0.38) | 0.24 | 0.11 | (0.02 | 0.02) | -1.06 | 3.10 | (-7.14 | 5.02) |
| 4 | 22 | 94.84 | 0.55 | 0.25 | (0.07 | 1.03) | 0.15 | 0.13 | (-0.10 | 0.41) | -0.01 | 0.17 | (-0.33 | -0.33) | 5.33 | 4.31 | (-3.13 | 13.78) |
| 5 | 22 | 79.34 | -0.13 | 0.26 | (-0.63 | 0.38) | -0.29 | 0.13 | (-0.55 | -0.03) | -0.08 | 0.19 | (-0.46 | -0.46) | -0.29 | 6.65 | (-13.32 | 12.74) |
| 6 | 23 | 62.38 | 0.47 | 0.20 | (0.07 | 0.87) | 0.34 | 0.15 | (0.04 | 0.64) | 0.27 | 0.20 | (-0.12 | -0.12) | -8.19 | 5.12 | (-18.23 | 1.84) |
| 7 | 22 | 48.04 | 0.39 | 0.23 | (-0.07 | 0.85) | 0.15 | 0.18 | (-0.21 | 0.51) | 0.33 | 0.29 | (-0.23 | -0.23) | 3.01 | 9.73 | (-16.06 | 22.07) |
| 8 | 22 | 42.08 | 0.40 | 0.14 | (0.12 | 0.68) | 0.28 | 0.17 | (-0.06 | 0.62) | -0.07 | 0.32 | (-0.70 | -0.70) | -5.71 | 5.29 | (-16.06 | 4.65) |
| 9 | 22 | 39.34 | 0.24 | 0.23 | (-0.20 | 0.69) | 0.03 | 0.19 | (-0.34 | 0.40) | 0.03 | 0.32 | (-0.59 | -0.59) | 18.89 | 10.28 | (-1.26 | 39.04) |
| 10 | 23 | 33.73 | 0.83 | 0.19 | (0.46 | 1.20) | 0.69 | 0.20 | (0.30 | 1.07) | 0.64 | 0.36 | (-0.06 | -0.06) | 9.44 | 4.73 | (0.16 | 18.71) |
| Cumulative estimates | | |  |  |  |  |  |  |  |  |  |  |  |  |  |  |  |  |
| 1 | 23 | 1456.67 | 0.04 | 0.07 | (-0.10 | 0.17) | 0.03 | 0.04 | (-0.05 | 0.11) | 0.10 | 0.03 | (0.05 | 0.05) | 0.33 | 0.12 | (0.10 | 0.56) |
| 2 | 45 | 819.67 | 0.05 | 0.06 | (-0.07 | 0.16) | 0.04 | 0.04 | (-0.03 | 0.11) | 0.10 | 0.02 | (0.06 | 0.06) | 0.11 | 0.09 | (-0.07 | 0.30) |
| 3 | 67 | 594.44 | 0.06 | 0.05 | (-0.04 | 0.17) | 0.06 | 0.04 | (-0.01 | 0.14) | 0.11 | 0.02 | (0.06 | 0.06) | 0.06 | 0.08 | (-0.09 | 0.22) |
| 4 | 89 | 474.29 | 0.09 | 0.05 | (-0.02 | 0.20) | 0.05 | 0.04 | (-0.03 | 0.12) | 0.11 | 0.02 | (0.07 | 0.07) | 0.02 | 0.08 | (-0.13 | 0.18) |
| 5 | 111 | 398.39 | 0.08 | 0.05 | (-0.02 | 0.19) | 0.04 | 0.04 | (-0.03 | 0.12) | 0.11 | 0.02 | (0.07 | 0.07) | 0.05 | 0.08 | (-0.10 | 0.20) |
| 6 | 134 | 342.41 | 0.09 | 0.05 | (0.00 | 0.19) | 0.03 | 0.04 | (-0.04 | 0.11) | 0.11 | 0.02 | (0.07 | 0.07) | 0.03 | 0.07 | (-0.11 | 0.18) |
| 7 | 156 | 302.01 | 0.10 | 0.05 | (0.01 | 0.20) | 0.04 | 0.04 | (-0.03 | 0.12) | 0.11 | 0.02 | (0.06 | 0.06) | 0.03 | 0.07 | (-0.11 | 0.16) |
| 8 | 178 | 270.76 | 0.11 | 0.05 | (0.02 | 0.20) | 0.06 | 0.04 | (-0.01 | 0.13) | 0.11 | 0.02 | (0.06 | 0.06) | 0.03 | 0.06 | (-0.10 | 0.15) |
| 9 | 200 | 246.05 | 0.11 | 0.04 | (0.03 | 0.20) | 0.06 | 0.04 | (-0.02 | 0.13) | 0.11 | 0.02 | (0.06 | 0.06) | 0.03 | 0.06 | (-0.09 | 0.15) |
| 10 | 223 | 224.76 | 0.12 | 0.04 | (0.04 | 0.21) | 0.09 | 0.04 | (0.01 | 0.16) | 0.11 | 0.02 | (0.06 | 0.06) | 0.02 | 0.06 | (-0.10 | 0.13) |

***Supplementary Table 8 – MR CAUSE results for estimation of the effect of CRP on Type 2 Diabetes***

|  | **(1)**  **MR CAUSE: r^2^=0.01, p=0.001** | | | **(2)**  **MR CAUSE: r^2^=0.1, p=0.001** | | |
| --- | --- | --- | --- | --- | --- | --- |
| ρ | 0.18  502 | | | 0.18  1013 | | |
| No. of SNPs |  |  |  |  |  |  |
| **Model comparison:** | **Z** | **p-value** | | **Z** | **p-value** | |
| Null vs Sharing | -1.00 | 0.16 | | -0.70 | 0.24 | |
| Null vs Causal | -1.19 | 0.12 | | -0.72 | 0.24 | |
| Sharing vs Causal | -1.08 | 0.14 | | -0.59 | 0.28 | |
| **Model parameters:** | **Median** | **95% CI** | | **Median** | **95% CI** | |
| Sharing model: eta | 0.67 | (0.05 | 3.37) | 0.30 | (-0.25 | 1.53) |
| Sharing model: q | 0.09 | (0.01 | 0.37) | 0.08 | (0.00 | 0.29) |
| Causal model: gamma | 0.08 | (0.01 | 0.15) | 0.04 | (0.00 | 0.09) |
| Causal model: eta | 0.86 | (-1.03 | 3.93) | 0.23 | (-0.93 | 2.80) |
| Causal model: q | 0.03 | (0.00 | 0.22) | 0.03 | (0.00 | 0.22) |

*Results from MR Causal Analysis Using Summary Effect estimates (MR CAUSE) estimation of the causal effect of CRP on T2D. Genetic variants are selected via LD pruning with a p-value threshold of 0.001 and an r^2^ threshold of 0.01 (column 1) or 0.1 (column 2). ρ = estimated correlation in summary statistics due to overlapping GWAS samples. No. of SNPs = number of SNPs which were selected via LD pruning and included in the CAUSE estimation. CAUSE estimation compares three models based on their expected log pointwise posterior density (ELPD): 1) a null model (no causal effect and no correlated pleiotropy), 2) a sharing model (no causal effect, but allows for correlated pleiotropy), (3) a causal model (causal effect and allows for correlated pleiotropy). Z statistics and p-values are shown for each pairwise model comparison, with a positive (negative) Z statistic indicating better fit of the former (latter) model. Posterior medians and 95% credible intervals are reported for the parameters of the sharing and causal model: eta = effect of shared factor on outcome, q = proportion of variants exhibiting correlated pleiotropy, gamma = effect of exposure (CRP) on outcome (T2D).*

***Supplementary Table 9 – MR-cML results for estimation of the effect of CRP on Type 2 Diabetes***

|  | **(1)**  *ρ=0.04  based on bivariate LD-score  regression intercept* | | | **(2)**  *ρ=0.05 based on phenotypic correlation  and GWAS sample overlap* | | |
| --- | --- | --- | --- | --- | --- | --- |
|  | ***theta*** | ***Std. Err*** | ***P-value*** | ***theta*** | ***Std. Err*** | ***P-value*** |
| cML-BIC | 0.09 | 0.02 | 3.08E-6 | 0.09 | 0.02 | 3.44E-6 |
| cML-MA-BIC | 0.09 | 0.02 | 6.82E-6 | 0.09 | 0.02 | 7.61E-6 |
| cML-BIC-DP | 0.11 | 0.07 | 0.09 | 0.11 | 0.06 | 0.08 |
| cML-MA-BIC-DP | 0.11 | 0.07 | 0.09 | 0.11 | 0.06 | 0.07 |

*Results from Mendelian Randomization with constrained Maximum Likelihood (MR-cML) estimation of the causal effect of CRP on T2D. The correlation in summary statistics due to overlapping GWAS samples was estimated using two methods: (1) the intercept in a bivariate LD-score regression (LDSC). (2)* $\rho=\frac{N_{0}}{\sqrt{N_{1}N_{2}}}r\left( x,y \right)$ *based on the size of the sample overlap* $N_{0}$*=356,339 between the two GWAS samples (*$N_{1}$*=575,531 ,* $N_{2}$*=490,089) and the phenotypic correlation* $r(x,y)$*=* *0.074*$.$ *Both GWAS samples included individuals from the UK Biobank (UKB), thus we assume the overlap to be equivalent to the smaller of the two UKB subsamples and estimate the phenotypic correlation in the UKB. Estimates of the causal effect theta of exposure (CRP) on outcome (T2D), its standard error and p-value are shown for four MR-cML estimators: cML-BIC=MR-cML with number of invalid SNPs k selected using Bayesian information criterion (BIC), cML-MA-BIC=MR-cML with weighted model averaging across all values of k, cML-BIC-DP=data perturbation version of cML-BIC, cML-MA-BIC-DP=data perturbation version of cML-MA-BIC.*

1.	Burgess, S., A. Butterworth, and S.G. Thompson, *Mendelian Randomization Analysis With Multiple Genetic Variants Using Summarized Data.* Genetic Epidemiology, 2013. **37**(7): p. 658-665.

2.	Bowden, J., G. Davey Smith, and S. Burgess, *Mendelian randomization with invalid instruments: effect estimation and bias detection through Egger regression.* International journal of epidemiology, 2015. **44**(2): p. 512-525.

3.	Bowden, J., et al., *Consistent estimation in Mendelian randomization with some invalid instruments using a weighted median estimator.* Genetic epidemiology, 2016. **40**(4): p. 304-314.

4.	Hartwig, F.P., G. Davey Smith, and J. Bowden, *Robust inference in summary data Mendelian randomization via the zero modal pleiotropy assumption.* International journal of epidemiology, 2017. **46**(6): p. 1985-1998.

5.	Burgess, S., F. Dudbridge, and S.G. Thompson, *Re:“Multivariable Mendelian randomization: the use of pleiotropic genetic variants to estimate causal effects”.* American journal of epidemiology, 2015. **181**(4): p. 290-291.

6.	Sanderson, E., et al., *An examination of multivariable Mendelian randomization in the single-sample and two-sample summary data settings.* International journal of epidemiology, 2019. **48**(3): p. 713-727.

7.	Hemani, G., K. Tilling, and G.D. Smith, *Orienting the causal relationship between imprecisely measured traits using GWAS summary data.* PLoS genetics, 2017. **13**(11): p. e1007081.
